# Modeling the population effects of epitope specific escape mutations in SARS-CoV-2 to guide vaccination strategies

**DOI:** 10.1101/2021.01.19.21250114

**Authors:** James S. Koopman, Carl P. Simon, Wayne M. Getz, Richard Salter

## Abstract

Escape mutations (EM) to SARS-Cov-2 have been detected and are spreading. Vaccines may need adjustment to respond to these or future mutations. We designed a population level model integrating both waning immunity and EM. We also designed a set of criteria for elaborating and fitting this model to cross-neutralization and other data in a manner that minimizes vaccine decision errors. We formulated four model variations. These define criteria for which prior infections provide immunity that can be escaped. They also specify different sequences where one EM follows another. At all reasonable parameter values, these model variations led to patterns where: 1) EM were rare in the first epidemic, 2) rebound epidemics after the first epidemic were accelerated more by increasing drifting than by increasing waning (with some exceptions), 3) the long term endemic level of infection was determined mostly by waning rates with small effects of the drifting parameter, 4) EM caused loss of vaccine effectiveness and under some conditions, vaccines induced EM that caused higher levels of infection with vaccines than without them. The differences and similarities across the four models suggest paths for developing models specifying the epitopes where EM act. This model is a base on which to construct epitope specific evolutionary models using new high-throughput assay data from population samples to guide vaccine decisions.

**Highlights:**

1. This model is the first to integrate both antigenic drifting from escape mutations and immunity waning in continuous time.
2. Tiny amounts of only waning or only escape mutation drifting have small or no effects. Together, they have large effects.
3. There are no or few escape mutations during the first epidemic peak and no effect of drifting parameters on the size of that wave.
4. After the first epidemic peak, escape mutations accumulate rapidly. They increase with increases in waning rates and with increases in the drifting rate. Escape mutations then amplify other escape mutations since these raise the frequency of reinfections.
5. Escape mutations can completely negate the effects of vaccines and even lead to more infections with vaccination than without, especially at very low waning rates.
6. The model generates population level cross-neutralization patterns that enable the model to be fitted to population level serological data.
7. The model can be modified to use laboratory data that determine the epitope specific effects of mutations on ACE2 attachment strength or escape from antibody effects.
8. The model, although currently unable to predict the effects of escape mutations in the real world, opens up a path that can guide model incorporation of molecularly studied escape mutations and improve predictive value. We describe that path.
9. Model analysis indicates that vaccine trials and serological surveys are needed now to detect the effects of epitope specific escape mutations that could cause the loss of vaccine efficacy.

## The nature of the problem that we address

SARS-CoV-2 has generated the most devastating pandemic in a century. Many aspects of SARS-CoV-2 dynamics remain unknown, including the risks of and reasons for reinfection. Reinfection by endemic coronaviruses is common in all age groups (Monto, DeJonge et al. 2020, Nickbakhsh, Ho et al. 2020) and SARS-CoV-2 reinfections are increasingly being documented (Babiker, Marvil et al. 2020). Reinfections might arise either because of 1) waning, in which host immunity wanes after recovery from infection, or 2) drifting, in which the virus evolves to escape immunity stimulated by prior infections, immunotherapies, or vaccines. Drifting can arise in many ways, including immunity-escaping mutations, positive selection that increases transmissibility without escaping immunity, and of course, randomness. In this paper, our use of the term drifting will always imply selective drifting due to escape mutations, although our model can capture increased transmissibility mutations as well.

The endemic coronaviruses might indicate what we can expect from SARS-CoV-2. In this vein, high variation in endemic coronavirus genomes at attachment sites led (Andersen, Rambaut et al. 2020) to suggest that virus variation might explain some reinfections. The genetic analyses of (Kistler and Bedford 2020) provided evidence that “…OC43 and 229E, are undergoing adaptive evolution in regions of the viral spike protein that are exposed to human humoral immunity. This suggests that reinfection may be due, in part, to positively-selected genetic changes in these viruses that enable them to escape recognition by the immune system.” This observation is consistent with patterns of genetic change in East Asian populations which suggest that among the 8 identified OC43 genotypes, genotypes vary enough to replace each other at different times (Lau, Lee et al. 2011) (Zhang, Li et al. 2015) (Zhu, Li et al. 2018). Recent cross-neutralization assays of genetic changes in the alpha coronavirus 229E during the 1980’s and 1990’s have shown clear genetic escape from immunity across time (Eguia, Crawford et al. 2020).

Studies of SARS-CoV-2 itself have indicated how escape mutations can become a problem. (Greaney, Starr et al. 2020) demonstrated the potential for extensive SARS-CoV-2 escape from monoclonal antibodies by mutating every nucleotide in the receptor binding domain (RBD). (Weisblum, Schmidt et al. 2020) also documented mutations that escape from monoclonal antibodies and noted that such mutations are present at low levels in circulating SARS-CoV-2 populations.

The first well documented genetic change associated with high SARS-CoV-2 transmission involved a D614G mutation (Hou, Chiba et al. 2020). This mutation has been well studied. It is not clear whether this is an escape mutation, even though the 614G variant is associated with higher viral load and younger age of patients (Volz, Hill et al. 2021). Evidence against this being an escape mutation and in favor of it generating greater contagiousness in other ways comes from neutralization assays. The original 614D variant is neutralized at a higher level in animals or humans whose immunity was stimulated by the 614G variant (Weissman, Alameh et al. 2020).

A second realization of the potential for escape mutations has emerged in South Africa. (Tegally, Wilkinson et al. 2021) documented a new variant with eight lineage defining mutations that has rapidly replaced the original pandemic strains. Two sites in the RBD of this new variant appeared to be under diversifying positive selection and are likely to represent escape mutations.

A third case has emerged in England. The new lineage (B.1.1.7) that came to dominate transmission in SW England is highly mutated and fits into an evolutionary pattern that suggests it emerged via extended evolution in an individual due either to immunodeficiency or to serum treatment of a case (Rambaut, Loman et al. 2020) (Volz, Mishra et al. 2020). The series of genetic changes in this virus have increased transmission by 50-70% with greater effects in the under 19 age group possibly indicating that some escape mutations are to endemic coronavirus epitopes (Davies, Barnard et al. 2021). A case of reinfection with this new escape mutant has already been detected (Harrington, Kele et al. 2021)

These situations call for an intensive surveillance and control effort. We need to detect a possible threat to public health from escape mutations before epidemics of escape mutant viruses spread widely. After all, it takes time to develop new vaccine stockpiles even if mRNA manufacturing is faster than older approaches. Vaccine conformation decisions need methods that distinguish rare escape mutations that will spread widely from those that will not. Knowledge of microbial behavior at a biological level in individuals is not enough for this task. We need knowledge of the population conditions and processes that generate population threats. Such knowledge needs to integrate theory about population processes with data that capture the effects of those processes. In other words, we need models that can be fitted to population level data.

## How we addressed this problem

Given the desired model properties stated above, we initiated a new approach to modeling the population level dynamics of escape mutations. In our model, we focus on immunity dynamics related to one or three epitopes. In reality there are many more. But their number is countable and manageable. We treat our models as if the epitopes we model are the only ones affecting immunity. This amplifies vaccine and natural infection immunity effects. Thus, in their current forms the models examined are not immediately applicable to vaccine decisions like those concerned with whether the B.1.1.7 variant requires a vaccine change. But the analyses we present create a platform for constructing more detailed and realistic epitope specific models that can be informed by the proliferation of epitope specific population level data, as they become available.

We modeled escape mutations as arising during transmissions to individuals who already have some immunity from which the virus can escape. Immunity in a person being reinfected (the infectee) leads to escape mutations in the virus coming from a source case (the infector). We assumed that there was a common drifting scale between infectors and infectees. In other words, we put both the antigenicity of SARS-CoV-2 and the immune capacity of someone previously infected with the virus on the same scale. The infectee has the same position on the drifting scale as their last infection.

To be more precise, we assume *M+*1 variants of the virus that can arise via escape mutations. For ease of exposition, we will often call these variants “drift levels” or sometimes simply “strains.” We write *I*(*h*), sometimes *I*_*h*_, for those currently infected with drift level *h*. We will usually reserve *h=*0 for the strain that initially infects the population. Currently uninfected, susceptible individuals are in two sets: *S* for those never before infected and *R*(*j,k*) for those recovered from a previous infection whose last infection was by drift level *j* and whose current level of immunity to strain *j* is *k*. The second subscript in *R*_*h*0_ represents how much the recovered individual’s immune system has waned from its maximum effectiveness. As their immune system wanes, the individual newly recovered from drift level *h* moves from *R*_*h*0_ to *R*_*h*1_ to *R*_*h*2_ and eventually to *R*_*hP*_ at constant rate *w*. So, for each drift level *h*, there are *P+*1 waning states *R*_*hk*_, *k*=0,1,…,*P*.

In this paper we consider three simple configurations of drift levels:

1. the *M+*1 drift levels lie at the integer points on a line, with extremes at *h*=0 and *h=M*;
2. the drift levels lie on a circle, like hour marks on a clock face (the circle is formed by tying the ends of the interval in configuration 1) so that both level 1 and level *M*-1 are equally adjacent to level 0 so this configuration has no extreme levels).
3. drifting occurs in patterns generated by γdichotomous alleles so that each drift level can now be written as a string of γcapital letters and small letters (or 0s and 1s), where such alleles cause the shape of drifting relationships to emerge as a γ-dimensional hypercube.

See Figure S1 in the Supplementary Material (SM). All three of these configurations have a natural notion of adjoining drift levels and the “edges” that join them. In this paper we measure the distance between two drift levels as the least number of edges that connect one to the other.

The question emerges: “What is the distance and direction in the drift space between infector and infectee where immunity in the infectee can cause the virus in the infector to drift one level?” For the linear configuration 1), we examine two situations: 1a) the influence can be from anywhere along the linear scale, 1b) the influence generating drifting only occurs when the infectee is at most one step away from the infector. In either case, the influence for drifting only occurs in one direction. Upon transmission, those viruses that escape a new host’s immunity are more likely to cause infection. For example, under our assumption that drift levels lie at the integer points on a line, if an individual in *I*(2) infects an individual in *R*(3,*k*), the virus in the infector could transit to *I*(1) or *I*(3) upon transmission. However, the individual in *R*(3,*k*) has more immunity to strain 3 than to strain 1. So, if there is a transit, it will be away from strain 3 to strain 1. More generally, when the infectee is higher along the scale than the infector, drifting will move one step lower along the scale. When the infectee is lower along the scale than the infector the drifting will move one step higher. This corresponds to drifting only being possible in a direction that escapes immunity in the infectee. Once variants that escape immunity are in an infectee with that immunity, the escape variant virus will have a growth advantage that across hundreds of virus multiplications on the way to peak infection will lead to the dominance of the escape variant.

The four models we examined are thus: Model 1 (corresponding to assumption 1a), Model 2 (corresponding to assumption 1b), Model 3 (the circular model), and Model 4 (the allelic hypercube generating model). In terms of an epidemiological and immunological interpretation of our topological models, we speculate that when there are multiple escape mutations in a single epitope, our linear topology is the more appropriate abstraction. Six linear steps, as in our models, would be an unusually high number. In contrast the hypercube allelic relationships correspond to situations arising when different escape mutations arise in different epitopes.

The issues we address with these four models seem not to have been considered before in either a scientific theory or a mathematical model formulation. In fact, we found no model that included both immunological waning and escape mutation drifting in continuous time. (Andreasen, Lin et al. 1997) have formulated a model with waning and drifting across seasons, but their model does not capture escape mutations in continuous time.

## Mathematical formulation of the models

The model form we analyzed is an ordinary differential equation SIR model in a single homogenously mixing population. We present the explicit equations of our model in an appendix after the references. SIR models divide people into only three categories: Susceptible, Infectious, and Recovered (*SIR*). We stick with this simplification but, as discussed in the last section, we divide the *I* into *M*+1 different drifting levels of virus and we divide the R into both M different drift levels and *P*+1 different waning levels.

Our mathematical formulation has two key elements not found in other SIR models: a drifting tensor and a susceptibility function. The <u>drifting tensor</u> is a general formulation for specifying how the immunity state of the infectee and the drift level of the infector will lead to drifting. It is an (*M*+1)×(*M*+1)×(*M*+1) array whose (*h,j,h’*) entry gives the probability that when an infector in *I*(*h*) transmits to an infectee in some *R*(*j,k*) the virus will drift from level *h* to level *h’*. The drift matrices for our four models are presented in Tables S1-S4 in the SM.

The <u>susceptibility functio</u>n *Z*(*h,j,k*) specifies the probability of transmission when an infector in *I*(*h*) contacts a susceptible in *R*(*j,k*). Probability of transmission *Z*(*h,j,k*) rises more as the distance between the two strains *h* and *j* increases. It also rises more as the susceptible person’s immune system wanes (higher *k*). See equation (1) in the Appendix.

The usual SIR parameter values used in our simulations are provided in the Appendix. To initialize the epidemic, we introduced one infection per 1 million into a continuous population nominally scaled to present results per 1000 individuals. All time scales were set to a week. We worked with 7 drift levels in Models 1, 2, and 3. In Model 4, we assumed there were 3 independent dichotomous alleles, so that there were 2_3_ = 8 drift levels. The resulting behaviors that we describe seem independent of the number of drift levels in Models 1, 2, and 3 or the number of independent alleles in Model 4.

We varied the drift rate *D* and the waning rate *W* to understand better the interactions between drifting and waning on the behavior of the system.

Table S5 in the SM provides some intuition for the effects of varying *W*. For example, in Model 1, at waning rate *W*=0.1, it takes 57 weeks for half of those entering *R*_00_ to reach *R*_06_. For *W*=0.01, that process takes 11 years.

Our waning functions have no redundant immunity. That is to say, as soon as there is any waning, there is susceptibility. The six waning steps are of equal size. In the last waning state, the susceptibility of previously infected individuals is 6/7ths of a fully susceptible, never before infected individual.

Drifting is likewise divided into six drifting steps and 7 drift levels for Models 1 and 2. For Models 3 and 4, however, there are only 3 steps to the most distant drift state, so we double the size of immunity loss for each step while leaving the last state having 6/7ths of full susceptibility.

## Model behaviors observed across the four models: without vaccination

**<u>In the absence of vaccination</u>**, the behavior of the four different models had remarkable similarities. Figure 1 shows the behavior of each of the four models varying *D* for fixed *W*=0.01. (Figure S2 in the SM does the same for fixed *W*=0.1.) Figure 2 does the same for varying *W* for fixed *D*=0.01. At a basic reproduction number of 2 we observed the following:

**Figure 1.**
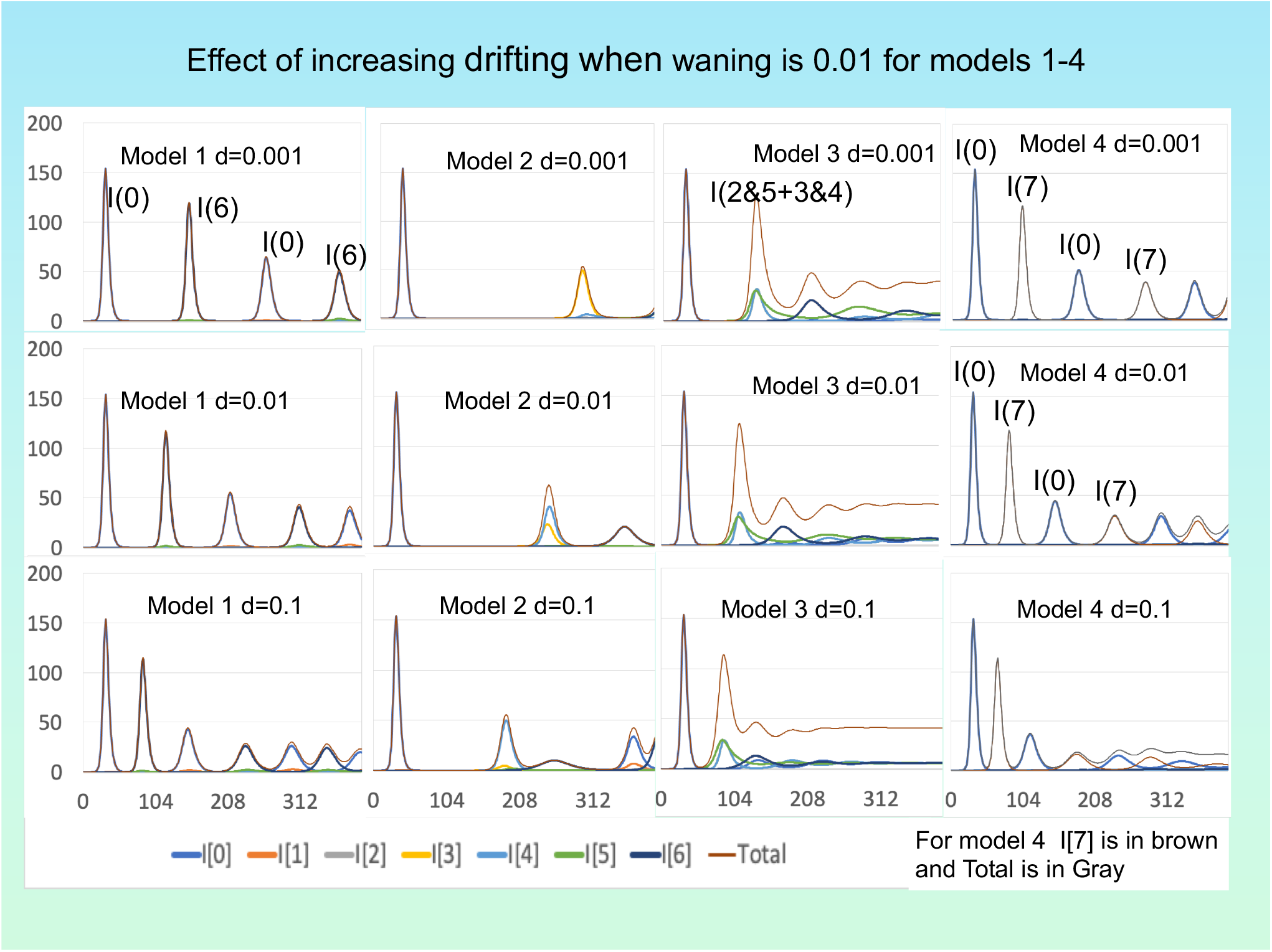
Runs of all 4 models with waning rate fixed at *W*=0.01 and drifting fraction varying at d=*D*= 0.001, 0.01, 0.1. In each panel the top curve gives the total number of infections with annotations for the dominant strains for different waves.

**Figure 2.**
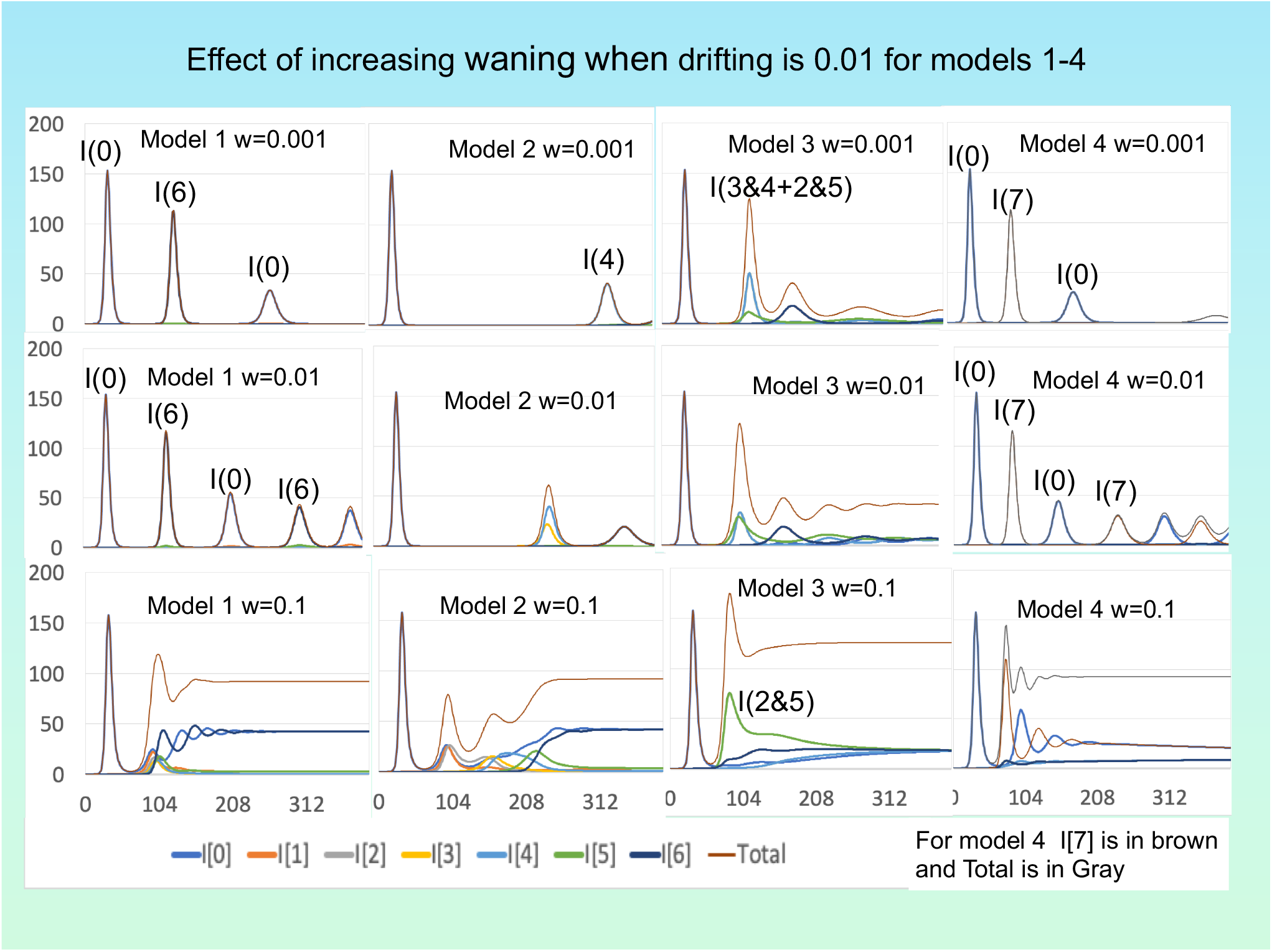
Analog of Figure 1, but now with drifting fraction fixed at *D*=0.01 and waning rate w*=W* varying at 0.001, 0.01, 0.1.

### Invariance of the first epidemic

The first epidemic is practically unchanged as *W* and *D* vary in the 3 graphs of Figures 1, 2 and S2, for Models 1 through 4. This invariance is expected since drifting in the model arises from reinfections, which are rare in the first epidemic; nearly all the infected are in *I*(0). The sharp rise and fall of the epidemic given homogeneous mixing contributes to this pattern. Perhaps the slower transmission expected from more realistic contact patterns would allow for some drifting during the first epidemic.

### Joint effects of waning and drifting

Without waning there was no drifting regardless of how high the drifting parameter was set. After the initial epidemic, with *W*=0 it took 78 years of new susceptible births before a second epidemic appeared. With tiny amounts of waning but no drifting (*W*=0.00001, *D*=0), results were similar. But with just an equally tiny amount of drifting (*W*=*D*=0.00001), the first rebound epidemic followed a few years after the first. So, at low levels of waning, the joint effects of waning and drifting are far greater than multiplicative. (See Figure S3 and S4 in SM.)

As seen in Figure 1 and the first two rows of Figure 2, for *W* and *D* values at or below 0.01, infections occur in discrete waves, with virtually no infection in between for Models 1, 2, and 4. However, with these values, Model 3 moves more quickly to continuous epidemics and then to equilibrium. There are two reasons for that. First, it takes only three drift steps instead of 6 to reach the maximal loss of immunity in Model 3. Second, each drift level has two different drift levels that are maximally distant from it. That means there are twice as many highly susceptible contacts to whom transmission can occur.

At *W* = 0.1, total infection does not remain at low levels in between waves but stays positive at the end of each epidemic and oscillates upwards in time, eventually reaching an equilibrium across all models. (Figures 2 and S2.)

As seen in Figures 1 and S2, increasing the drifting parameter *D* accelerates the occurrence of rebound epidemics after the first epidemic for waning rates of 0.01 and 0.1. It also accelerates the attainment of equilibrium levels of infection. Model 2, which has the lowest fraction of contacts that induce escape mutations, has the longest intervals to rebound epidemics and lowest infection rates in those epidemics. Model 3 has the same limitations on the fraction of contacts that induce escape mutations as Model 2. However, its circular form adds contacts between the ends that induce mutations as well as mutations from one end to the other. These additional drifting paths as well as the additional contacts that induce drifting at what are end levels in Model 2, result in Model 3 showing drifting comparable to what is seen in Model 1.

Increasing the waning rate accelerates the appearance of the first rebound epidemic in all four models, as Figure 3 illustrates for *D*=0.01 in Model 1. As Figure S4 in the SM illustrates, this acceleration is a little slower in Model 4.

**Figure 3.**
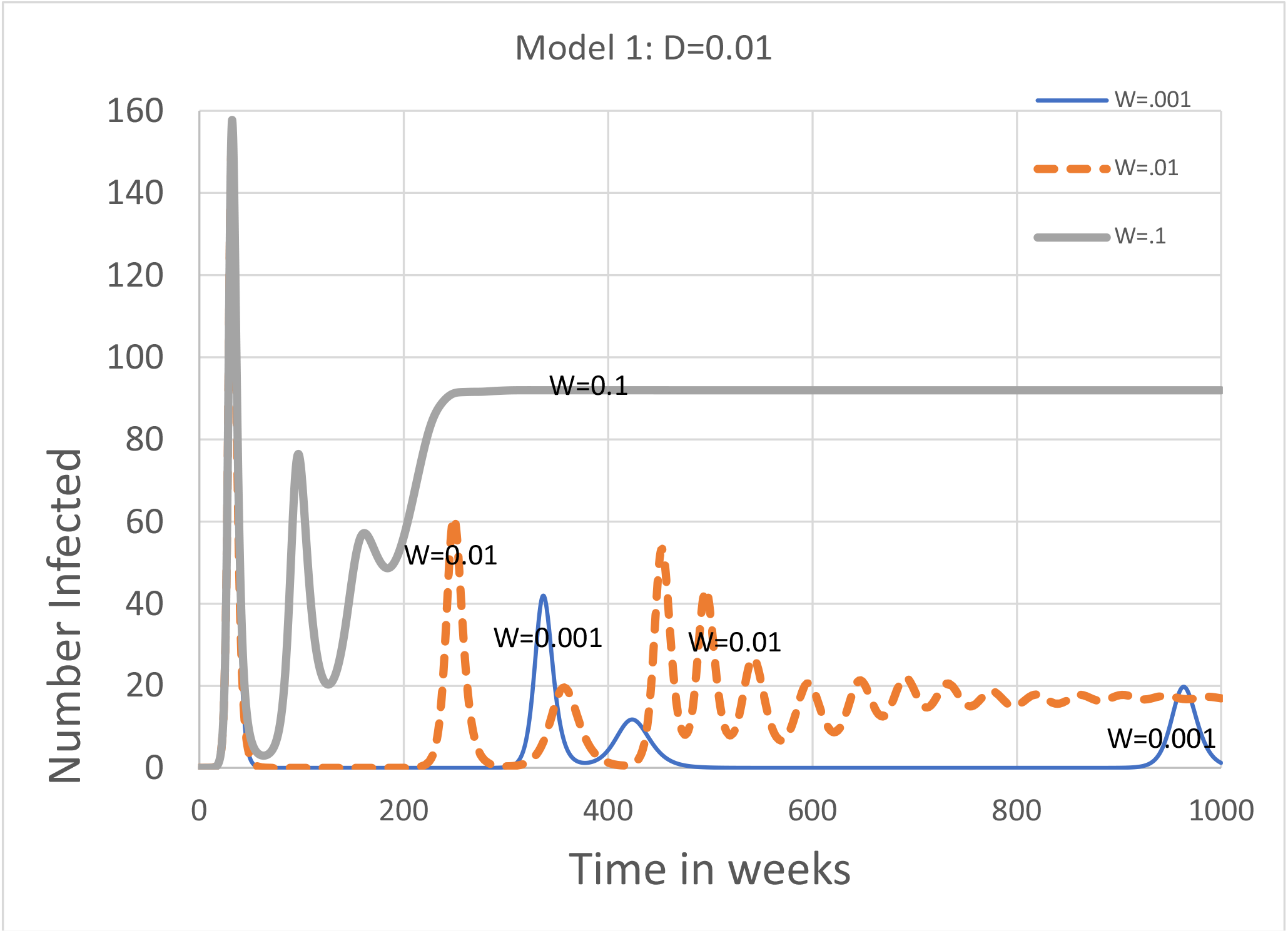
Simulation of Model 1 with drift fraction fixed at *D*=0.01. As waning rate *W* increases from 0.001 to 0.01 to 0.1, the time between waves shrinks and the level of infection increases noticeably.

### Equilibrium infection levels

Prevalence reaches an equilibrium in all four models, for all parameter values. In the 0.001 to 0.1 parameter ranges, endemic equilibrium infection levels are an increasing function of the waning rate *W* and are mostly unaffected by the drifting parameter *D*. As seen in Figure 4, the equilibrium infection levels were identical for models 1 and 2 and nearly identical for Model 4. But they were considerably higher for model 3; in model 3 each drift level has two different drift level that are maximally different from it while all of the other models each drift level has only one. (See Figures 4 and S1.)

**Figure 4.**
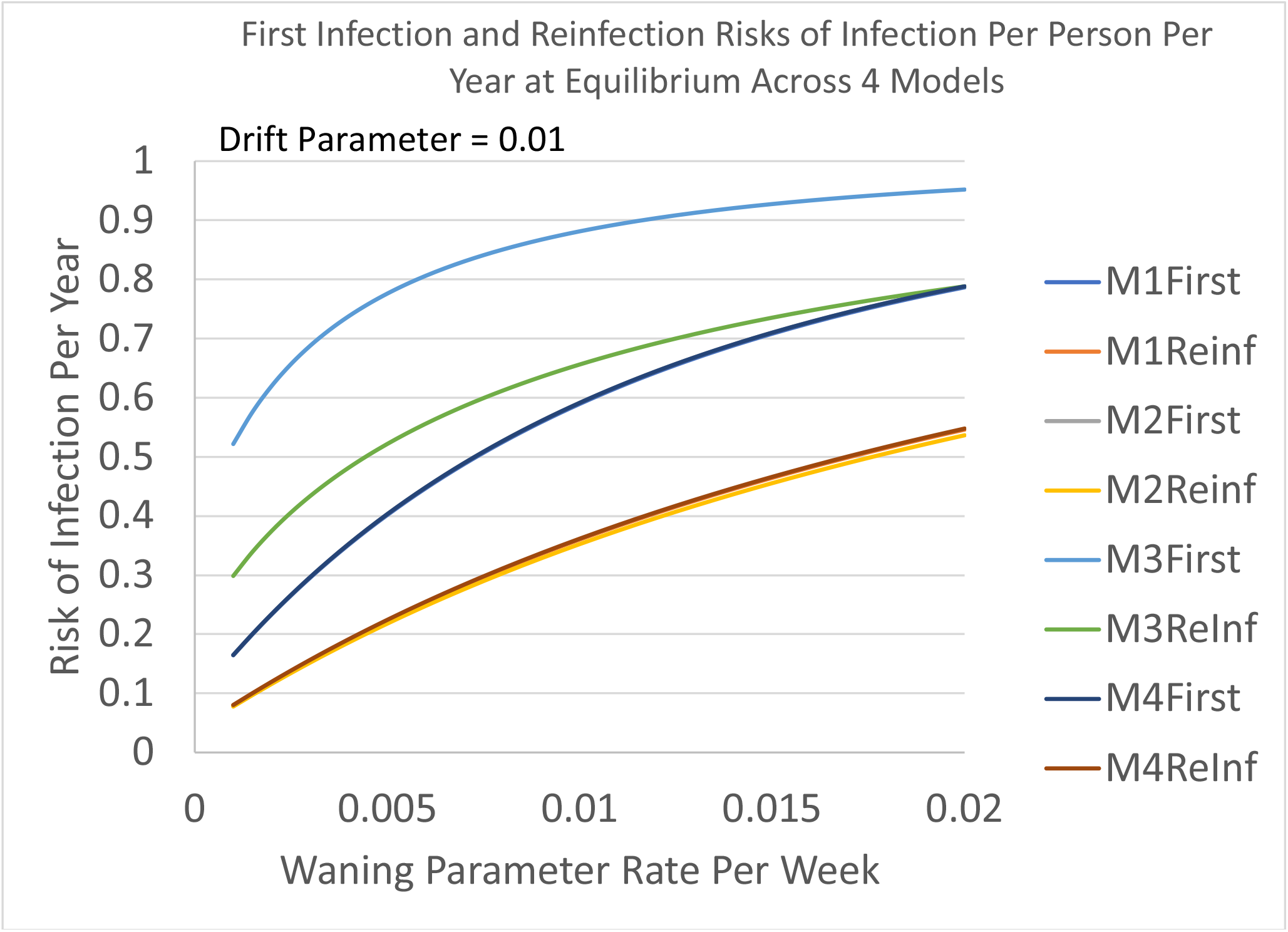
Graphs of the risk of first infection and risk of reinfection at equilibrium for all 4 models as the waning rate *W* increases. The top two graphs are for Model 3, the bottom two are for Models 1, 2, and 4. In each case, the risk of first infection is greater than the risk of reinfection.

The drifting parameters accelerate the attainment of equilibrium, but they do not increase equilibrium levels. In fact, for Models 1 and 2 they lower the equilibrium a little as they decrease the equilibrium accumulation of infections at the ends of the lines of drifting in these models. The equilibrium infection level is unchanged in Models 3 and 4 as the drifting rate changes.

The equilibrium infection levels had risks for reinfection that were comparable to the observed reinfection rates by endemic coronaviruses in older individuals when the waning rate was in the 0.001 to the 0.01 range. The true rates of reinfection, including asymptomatic infections and those mild enough to not be detected, are likely to correspond to a higher range. The endemic first infection risks were more than twice as high as the endemic reinfection risks (Monto, DeJonge et al. 2020, Nickbakhsh, Ho et al. 2020) in all models. (See Figure 4).

### Timing and composition of the subsequent epidemics

The drifting parameter has significant effects on both the timing of the first rebound epidemic and the extent of drifting in that epidemic. Except at very high waning rates and very low drifting rates, the first rebound epidemic – wave 2 -- has a virus drifted from the first epidemic.

As mentioned above, the first epidemic is dominated by the initial strain – level 0 in most of our simulations. Except in Model 2, the second wave is dominated by the drift level the furthest from level 0. In Model 1 the drift from strain 0 to strain 6 occurs rapidly during the end of the first epidemic. The drift restriction that distinguished Model 2 from Model 1 slows down this all-too-rapid drift so that intermediate drift levels dominate wave 2 in Model 2. Strains 0 and 6 take turns dominating alternate epidemics in Model 1, and eventually in Model 2.

Drift levels 3 and 4 dominate the second wave in Model 3, but soon thereafter all seven strains equilibrated to the same value. In the allelic Model 4, if we write ABC for the initial strain, then abc dominates the second wave, ABC dominates the third, then abc, until eventually all eight strains equilibrate to the same value.

## 6) Model behaviors observed across the four models: with vaccination

### Vaccination effects

were explored by beginning vaccination at the end of the first epidemic and keeping it constant thereafter. Vaccination rates of 0.25 and 1.0 per person per year were examined. The vaccine modeled generated exactly the same immunity as would an infection. The immunity is against infection. There is no immunity against disease given infection in this model. Specifically, vaccination moves a susceptible to state *R*(0,0), where drift level 0 is the initial infecting level – the one that thoroughly dominates the epidemic wave that precedes the introduction of the vaccine. The models discussed here have a small number of epitopes and no redundant immunity. That makes the insights generated by our analysis relevant for constructing more realistically detailed models, but less relevant for interpreting real world vaccine patterns.

When both the waning rate and drifting rate are at 0.1 as in Figure 5, we see for all models that vaccination has tiny but beneficial effects on the first rebound epidemic for models 1 and 4 while the effects for models 2 and 3 are small but not so negligible. The effects on endemic levels are more uniform across models; vaccination decreases infection prevalence at equilibrium.

**Figure 5.**
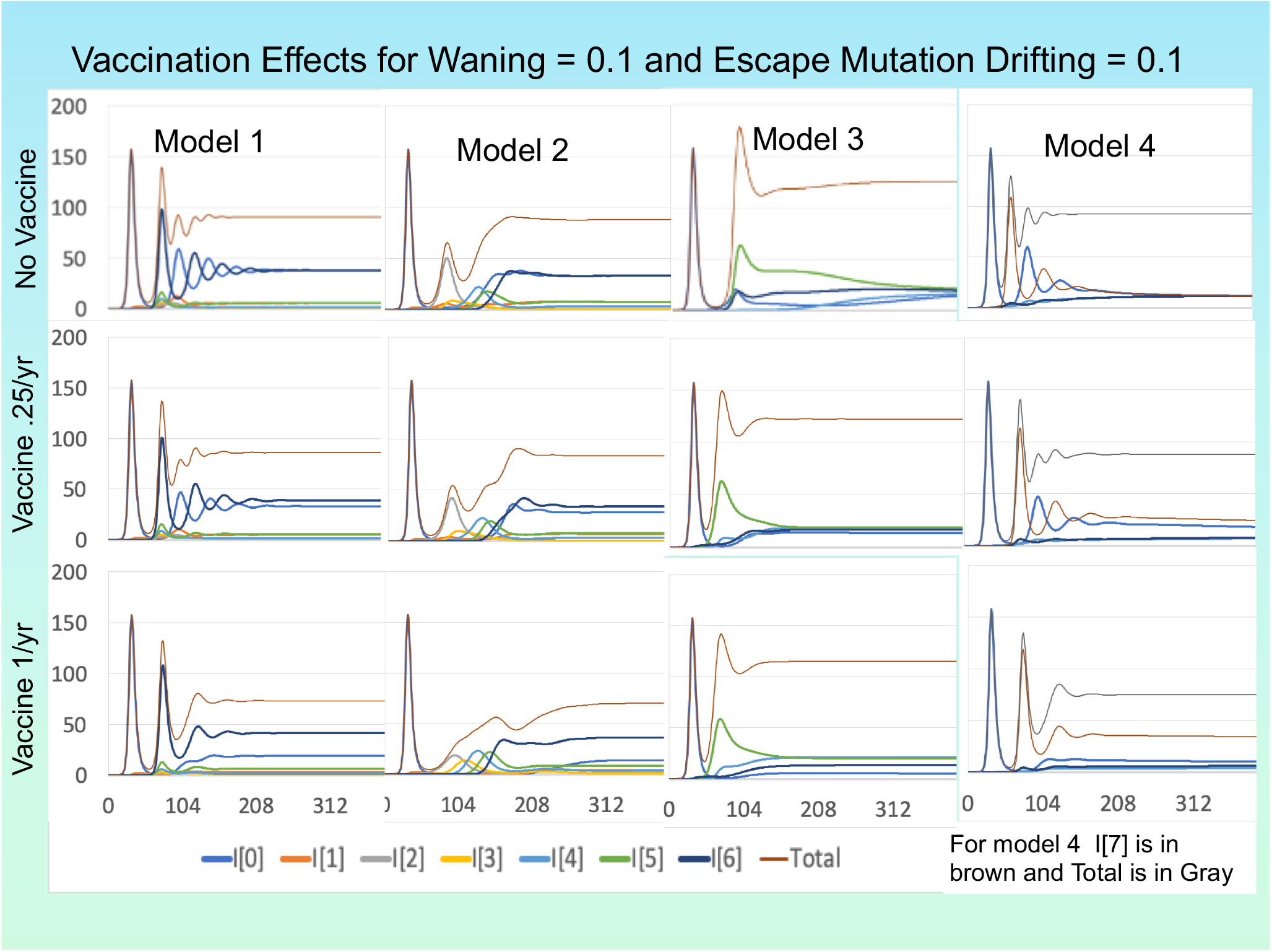
For each of the 4 models, *W* and *D* are fixed 0.1. The first row presents the graphs for the case of no vaccination; the second row presents partial vaccination (25% of the population per year); the third row presents full vaccination per year.

When the waning rate is decreased to 0.01 as in Figure 6, however, we see small but positive vaccine effects when the vaccination rate is only about a quarter of the population per year. However, when the vaccination rate goes up to 1 per year, vaccination has negative effects. It increases the frequency of rebound epidemics and raises the equilibrium levels of infection.

**Figure 6.**
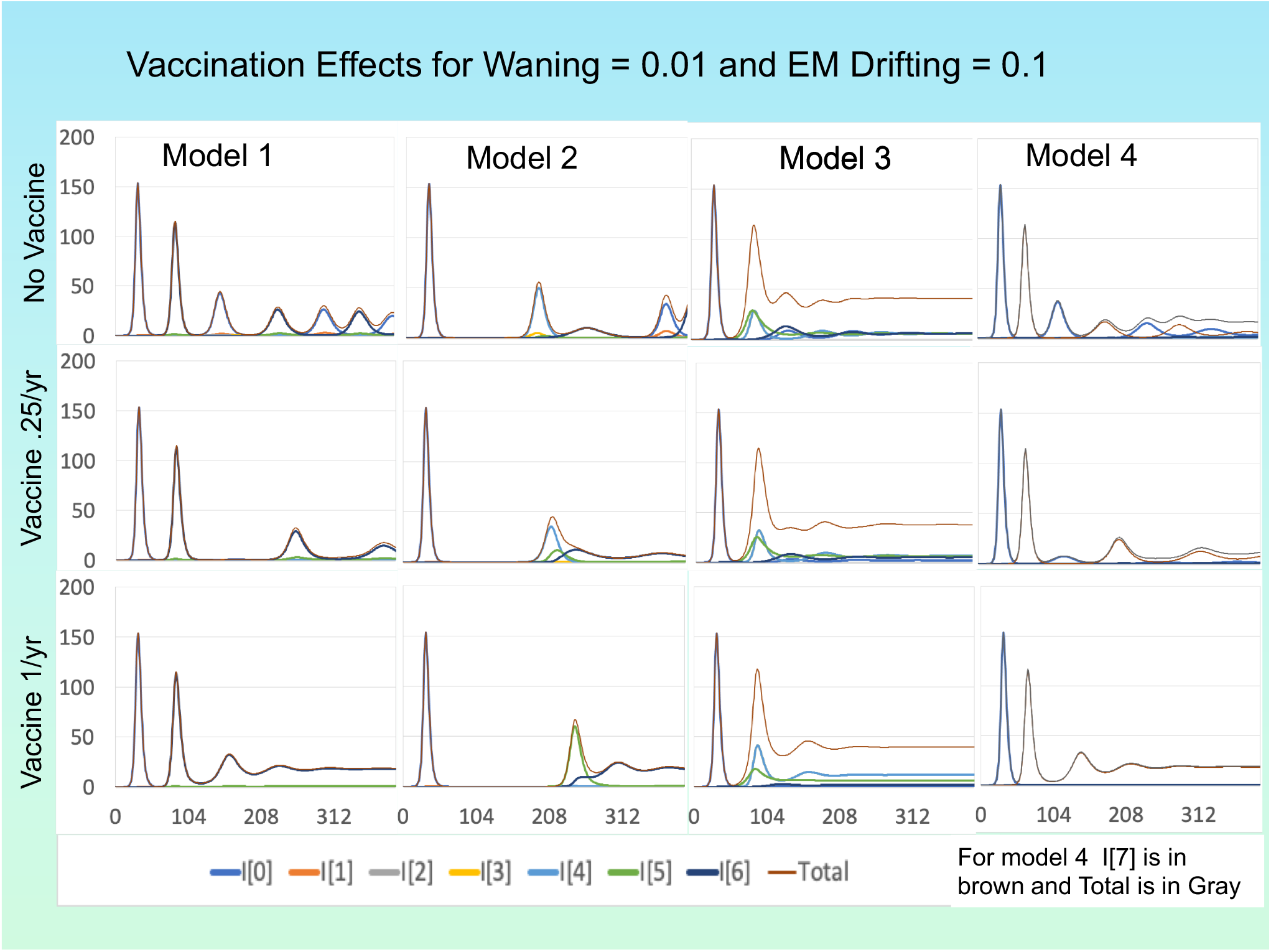
This is the analog of Figure 5, but with the waning rate *W* at .01 instead of 0.1. Full vaccination is more problematic here than in Figure 5.

Similar patterns are seen in Figures S4 and S5 where the drifting level is 0.01. At waning rate 0.1 vaccines reduce infection levels a little at both vaccination rates. At waning rate 0.01 there are slight beneficial effects of low vaccination rates, but harmful effects at the high vaccination rates with higher rates of infection with the vaccine than without it.

These harmful effects occur in all cases because high levels of vaccination eliminate strains with drift levels like those of the pandemic strain and the vaccine. They thus increase the frequency of individuals that have the highest levels of susceptibility to the most drifted strains. Those drifted strains can thus cause more infections than would be the case if the population were oscillating between the drifting extremes. Model 3 is less subject to these nefarious effects for the same reasons it stood out with regard to parameter effects when there was no vaccination. In model 3, the number of escaped strains that can take advantage of immunity levels stimulated by the vaccine or pandemic strain is greater so the vaccine has fewer individuals it can drive into the most drifted state.

Figures S6 and S7 make it easier to see the differences in vaccine effects when the waning rate changes from 0.01 to 0.1 by putting all three vaccination rates in the same figure.

## Generation of cross-neutralization assay and epitope specific serology results

Cross neutralization assays have long been used to determine whether new strains of viruses have developed escape mutations. For example, there has been great interest in the cross-neutralization assays of people infected in the U.S. for the new virus from SW England. But these assays have not previously been used to fit waning and drifting parameters of population models.

A strength of our model formulation is that it generates population patterns of immunity. As a result, our model can be fit to population level serology data and thus inform decisions about which epitope conformations to include in updated vaccines. Describing how cross neutralization tables are generated by the model and how they change over time given different parameter values illustrates the potential of fitted models to be used in vaccine composition decisions.

At a population level, cross-neutralization analyses are performed on a sample of individuals from a population. Each person provides a sample of antibody sera to assay the strength of their immune system to neutralize two or more viruses that may have been isolated at different times or from different populations. If the population patterns ascertained indicate that the titers of everyone are the same against each virus, then there is no drifting between different viruses. If there is a statistically significant difference, then the viruses have drifted to be different from each other.

To generate cross neutralization data from our models’ output, we use the *Z*(*h,j,k*) function as formulated by function (1.1) in our model (without *B*) to calculate the susceptibility of all individuals who have recovered from a previous infection. Since the *Z* function is a transmission parameter, it corresponds to the inverse of the neutralizing level. To make that number correspond to a titer, we divide the interval [0,1] into 10 equal subintervals that could correspond to 10 sequential dilutions of the sera for the neutralization assay. Both waning and drifting determine the neutralizing antibody levels according to equation (1.1). We present an example of such a construction in the Supplementary Material.

Comparison of model-generated cross-neutralization tables and corresponding tables from serology labs can be used to fit the model waning and drifting parameters. Such model fits can then be used to project how the immunity levels in the population will affect the spread of new virus variants with escape mutations. They can also be used to project how quickly further escape mutations might emerge. The validity of such projections requires more models that specify a series of specific potential escape mutations. The type of data generated by (Shrock, Fujimura et al. 2020) in combination with the type of data generated by (Greaney, Starr et al. 2020) and (Greaney, Loes et al. 2021) would be especially helpful in this regard.

There are too many epitopes and too wide a variety of immune responses to those epitopes for a model to capture all that information extensively enough to validate a vaccine decision. Because of this, deciding on the best way to validate a vaccine decision becomes a crucial issue that we deal with in the next section.

## Using the model and data to make decisions about vaccination: DRIA

The model we have presented is a first step in a process needed to make valid scientific or policy decisions. The Decision Robustness and Identifiability Analysis (DRIA) process we propose, as shown in the Figure 7 has a number of distinct steps that will make subsequent investigations more productive.

**Figure 7:**
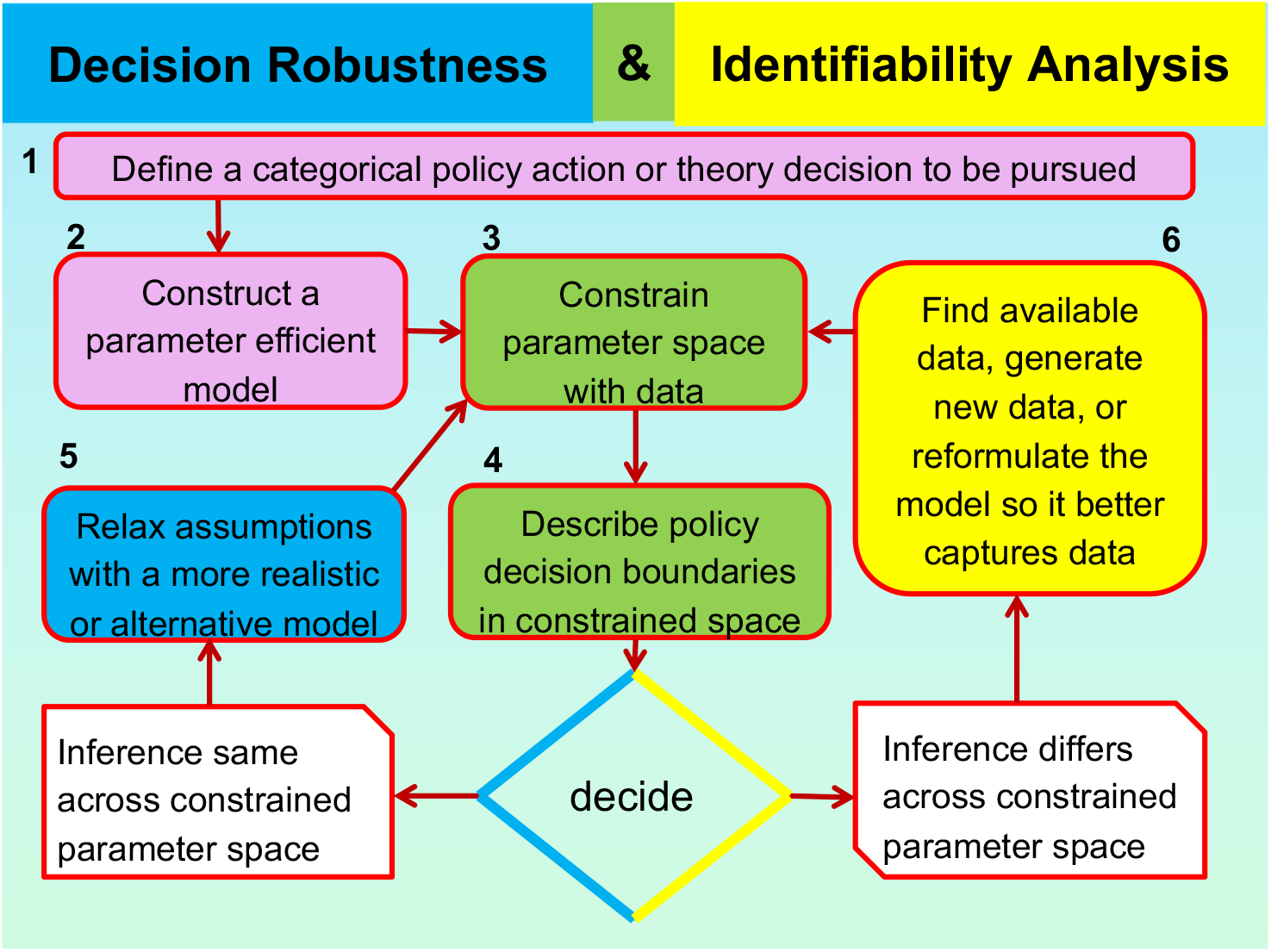
The DRIA algorithm steps

The enumerated steps are as follows:

1. Formulate the decision to be made in terms acceptable to policy makers. This requires a dialogue that has yet to take place. One such decision could be “All needed steps should be taken now so that SARS-CoV-2 vaccines can be updated in a timely manner to counteract escape mutations.”
2. Construct a simple model like the one in this paper that helps make the chosen decision.
3. Fit the model to data.
4. Assess whether a decision is identifiable given the data used. It is identifiable only if all of the parameter space that fits the data is consistent with only one decision. This requires two tasks to be completed. First, decision boundaries must be mapped out in parameter space. Second, the model must be fit to data and the parameter space consistent with the data must be mapped out. Some data to be used has been discussed above. This key step is addressed by using one of the modern approaches to fitting models to data (Funk and King 2019).
5. If the decision is identifiable, proceed with a decision robustness loop. That puts the decision on firmer grounds by reducing the possibility that unrealistic model assumptions could be determining the decision.
6. If the decision is not identifiable, proceed with a decision identifiability loop by seeking more informative data or better use of available data to improve decision identifiability.

DRIA loops address two major sources of errors when using models to make decisions about complex dynamic systems:

1. Robustness error: Some aspect of model structure that does not correspond to reality leads to a wrong decision.
2. Identifiability error: The fit of the model to data which leads to a decision is not the only fit possible and other fits could lead to a different decision.

If one is pursuing a decision about a scientific theory formulation, decision robustness and identifiability loops should go on infinitely. Policy decisions, in contrast, must be made quickly before things get out of control.

Instead of pursuing a model with all the detail that molecular biology can generate, we advocate taking all of that molecular biology into account to find abstractions where, step by step, one could relax simplifying assumptions to capture ever more of that molecular biology. Further thoughts on how to do this are included in the Supplementary Material.

## Needed steps for model development

### Improve Immune memory in the model

As attempts are made to pursue the strategy just outlined, we need to understand better how various aspects of our model formulation affect model transmission dynamics. A first issue to study is the effect of relaxing the assumption that reinfection wipes out prior immunity and leaves only the stamp of the last drifting level of infection - the assumption that our *R*(*j,k*) depends only the previous infection by strain j. One solution is to make immune memory specific for each drift state. It seems likely that if immunity to old strains is retained, the extreme swings which are exceptionally strong in Models 1 and 4 should be ameliorated. Later, as elaboration of the model in a DRIA context proceeds to specify immunity to specific epitopes, immune memory could be formulated as epitope specific. It might even specify antigen variation within epitopes. This would enable data being generated about epitope specific immunity to answer key questions in vaccinology by fitting our model to such data.

#### Formulate immunity effects more realistically

The model we analyzed had no redundant immunity which, even after some waning, still provides complete protection. It also had each drift level making a separate contribution to immunity which was removed by waning. A formulation with some redundant immunity that wanes quickly, then a somewhat slower waning followed by a longer term quite slow waning seems reasonable. Hopefully the numerous studies now underway to examine waning immunity will provide needed data. Our model will assist in separating out the waning from drifting in such data.

Both drifting and waning in our model are formulated as consisting of elements that independently add or subtract immunity from the total possible immunity. Once immunity to several drift levels comes into play, the necessity to formulate a joint effect for immunity arises. It takes extensive data to determine joint effects. Using the DRIA process should keep the joint effects that need to be determined to a minimum.

#### Mutation effects on contagiousness

In most of the experimental observations in (Greaney, Starr et al. 2020) escape mutations also led to decreased attachment to the ACE2 receptor. A simple solution there is to modify the overall effective contact rate parameter by a term specific for each drift level within the Z function of waning and drifting. That simple change would also enable modeling of changes like the D614G mutation that increases transmissibility without necessarily escaping pre-established immunity.

Efforts to fit this model to data, however, should not await all of the increased understanding discussed above. Focusing on making decisions regarding vaccine effect surveillance systems and vaccine modifications to counter escape mutations should guide which of the more theoretical issues are more intensely pursued.

## Conclusion

Recently a group of prominent infection transmission system modelers published an SIR model like ours in Science, but just with waning and not drifting (Saad-Roy, Wagner et al. 2020). They examined immune life history, vaccination, and the dynamics of SARS-CoV-2 over the next 5 years. When they took seasonality into account, they found that complex system phenomena could accelerate or increase the size of rebound epidemics and alter the effectiveness of vaccinations. While they did not model drifting, they acknowledged its importance and the key need to establish the impact of viral evolution, coinfection, and other pathogen characteristics on COVID-19 infection and disease. The findings in this paper reinforce that call.

The theory, data, and models needed to guide us to effective and long-lasting vaccine control of coronaviruses should rapidly go well beyond what is in this paper. The SARS-CoV-2 pandemic has led to the flourishing of diverse molecular methods that advance the science of vaccinology. These create the potential to specify the epitopes that stimulate immunity, the variation in immune responses to each of those epitopes, the classification of immune response components by their effectiveness for controlling infections, the prediction of what peptides will stimulate B and T cell responses in different people, the prediction of which immune responses might stimulate auto-immunity, and the prediction of the effects of mutation in each amino acid of a virus. None of these advances by themselves can indicate what escape mutations are likely to emerge de novo or how newly detected genetic variations will spread in different populations. To achieve these predictive goals, a model that builds on all the epitope specific advances listed above, but that does so judiciously in a manner that advances identifiability of decisions based on these advances is needed. The needed model is more elaborate than the model we have presented. But it can be built on the basis of that model.

## Data Availability

There is no data in this paper. It is purely a modeling study.

## APPENDIX: Mathematical formulation details

The total population *N* is partitioned into never-infected individuals in *S*, individuals currently infected with strain *h* in *I*_*h*_, and previously infected individuals in *R*_*jk*_ whose last infection was from strain *j* and who have immunity level *k*. Here, drift levels *h* and *j* go from 0 to *M*-1 and the immunity level *k* goes from 0 to *P*.

## Transmission and Waning

New infections can occur when an infected in state *I*_*h*_ meets a recovered individual in state *R*_*jk*_ (our compact notation for *R*(*h,j*)) *with* the probability of transmission increasing: 1) as the waning level *k* increases, and 2) with increases in the distance (in some metric) dist(*h,j*) between the drift level *h* of the infected and last former infection level *j* of the susceptible in *R*_*jk*_. For example, there is no transmission when an *I*_*h*_ encounters a *R*_*h*0_ and the highest probability of transmission when an *I*_0_ encounters someone in *R*_*hj*_. To quantify this probability of transmission between an *I*_*h*_ and an *R*_*jk*_, we combine the risk of infection due to waning 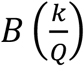 with the risk due to drifting 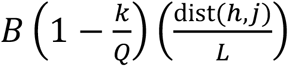. Since there are *P*+1 waning states and *P* waning steps, *Q* must be ≥*P*. In all four models 1 and 2 we use *Q*=*P*+1=7. Since there are *M*+1 drift levels and *M* drift steps, *L* must be ≥*M*. In models 1 and 2 where there are *M* = 6 drifting steps, we use *L=M*+1. In Models 3 and 4 where there are only *M* = 3 drift steps, we use *L* = 3.5 so that the ratio is the same between models. The more general values *Q* and *L* allow us to increase or decrease the relative weight of waning or drifting on transmission probability in our model – flexibility that we will use in the SM. The total probability of transmission when an infective in *I*_*j*_ contacts a susceptible in *R*_*hk*_ is:

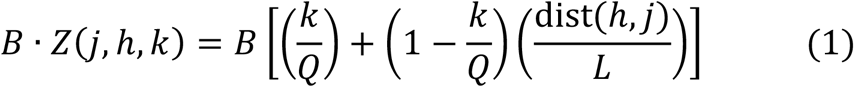

This formulates independent joint effects of waning and drifting on infection risk with drifting adding susceptibility beyond the current level of waning. It implies that drifting and waning operate on the same scale in terms of ability to increase susceptibility.

## Drift Tensors

For general spaces of drift levels, drift tensors (or matrices) present a formulation to model drifting. Suppose that a virus has *M+1* drifting levels. One can construct an (*M*+1)×(*M*+1)×(*M*+1) “drifting tensor” that captures drifting processes like ours. Consider the case where an individual in *I*_*h*_ (infected by a virus at level *h*) transmits the virus to a susceptible in *R*_*j*\*_ (most recently infected by the virus at level *j*). Let h’ denote a level to which the level *h* virus might drift at such a transmission. Let *ϕ*(*h,j,h’*) denote the probability that such a drift to level *h*’ occurs. These *ϕ*(*h,j,h’*)s are the entries of the drift tensor. See the examples in Tables S1-S4.

### Vaccination

In all the models in this paper, we assume that a vaccination program begins after the first wave and goes on continuously. Individuals in *S* or any of the *R*_*jk*_*’*s are vaccinated. A fraction *v* of these susceptibles is vaccinated each week; our default *ν* is 0.02 which is more than once per person per year. Unless otherwise specified, we assume that the vaccine enhances immunity only to strain 0, the strain that dominates the first wave. In particular, we assume that vaccinated susceptibles receive the same protection as if they had recovered completely from a strain 0 infection, i.e., they move into compartment *R*_00_.

The system of differential equations that describes our model for general drift spaces is as follows:

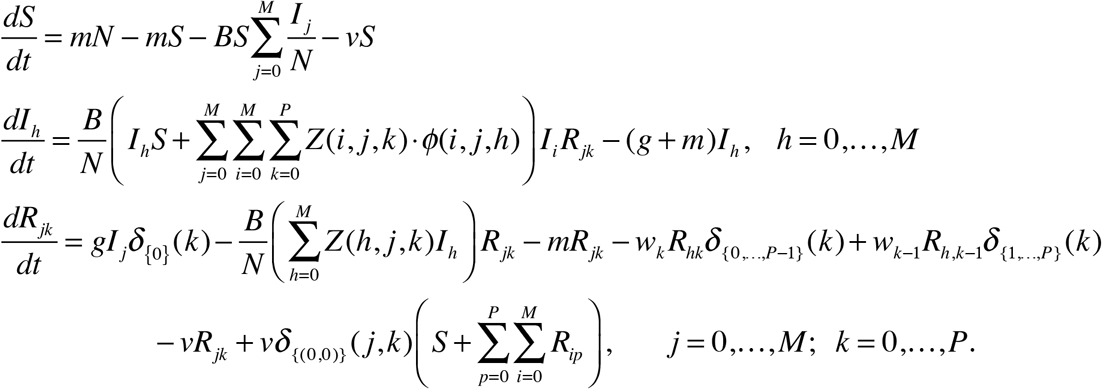

Here, for any subset Y of a set 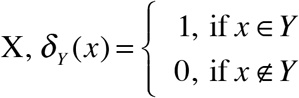.

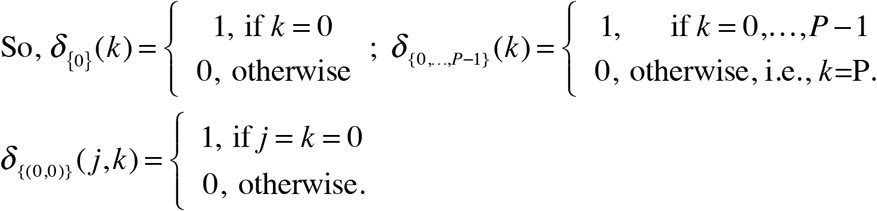

The first equation describes the infection and vaccination of never-infected susceptibles. The second set of equations describes the dynamics of those infected with drift level *h* – the *I*_*h*_’s, who recover at rate *g* and die at rate *m*. New *I*_*h*_’s arise when an *I*_*h*_ infects an *S*, infects any *R*_*jk*_ without drifting, or when an *I*_*i*_ drifts to an *I*_*h*_ upon infecting an *R*_*jk*_. The probability of the latter occurrence is the probability *Z(i,j,k)* that any given *I*_*i*_ will infect an *R*_*jk*_ times the probability *ϕ*(*i,j,h*) from the drift tensor that the *I*_*i*_ will drift to an *I*_*h*_ upon such an infection.

In the third set of equations, *R*_*jk*_’s increase when an *I*_*j*_ recovers or when an *R*_*jk*-1_ wanes. *R*_*jk*_’s decrease when they wane to an *R*_*jk*+1_ or upon vaccination. The last line in the above equations keeps track of the vaccination process when the newly vaccinated enter *R*_00_.

We simulated the model delineated in the Supporting Material using the Berkeley Madonna Software (Madonna 2021) always checking to see that the shortening the step size did not change the results. For the simulations in this report we set effective weekly transmission rate parameter *B*=1 or 1.5 and weekly rate of recovery *g=0*.*5*, so that the underlying basic reproduction number is *R*_0_=2 or 3. The birth and death rates were set at 1/(75×52). All time scales were set to a week. We introduced one infection per 10 million into a continuous population denoted as having size 1000. Numerical solution of the model used Runge-Kutta 4 and the stability of numerical solutions were evaluated across smaller time steps.

A Numerus Model Builder version of Model 2 was built independently of the Berkeley Madonna model. A web version of this Numerus model that includes a seasonal driver is under construction and revision.

## SUPPLEMENTARY MATERIAL

1. Drift Matrices for Models 1, 2, 3, and 4
2. More figures and tables for model behavior without vaccination
3. More figures and tables for model behavior with vaccination
4. Examples of model generated cross neutralization tables
5. Varying model parameters
6. DRIA informed vaccine decisions

## S1. Drift matrices for our models

### Drift Matrices

Drift matrices present a formulation to model drifting. Consider the case where an individual in I_h_ (infected by a virus at level h) transmits the virus to a susceptible in R_j*_ (most recently infected by the virus at level j). Let h’ denote a level to which the level h virus might drift at such a transmission. Let ϕ(h,j,h’) denote the probability that such a drift to level h’ occurs. These ϕ(h,j,h’)s are the entries of the drift matrix.

Table S1 presents the drift matrix for the drift process in Model 1 in which there are 7 drift levels. To represent 7×7×7 underlying drift matrix on this two-dimensional page, for every I_h_ and R_j*_, we list in row h and column j of Table S1 all the ordered pairs *(h’, ϕ(h,j,h’))* for which *ϕ(h,j,h’)≠0*. For example, in the third row, fourth column of Table S1, we see that when an I_2_ infects an R_3k_, there is probability D that the I_2_ will drift to an I_1_ and probability 1-D that there will be no drift and the newly infected will enter I_2_.

**Table S1.** Drift Matrix for Model 1, representing the 7×7×7 drift matrix for the model we simulate. The entry in row h and column k contains all the ordered pairs *(h’, ϕ(h,j,h’))* for which *ϕ(h,j,h’)≠0*.

| | $R_{j^*}$ | 0 | 1 | 2 | 3 | 4 | 5 | 6 |
| --- | --- | --- | --- | --- | --- | --- | --- | --- |
| $I_h$ | | | | | | | | |
| 0 |  | 0, 1-D<br>1, D | 0, 1 | 0, 1 | 0, 1 | 0, 1 | 0,1 | 0,1 |
| 1 |  | 1, 1-D<br>2, D | 0, D/2<br>1, 1-D<br>2, D/2 | 0, D<br>1, 1-D | 0, D<br>1, 1-D | 0, D<br>1, 1-D | 0, D<br>1, 1-D | 0, D<br>1, 1-D |
| 2 |  | 2, 1-D<br>3, D | 2, 1-D<br>3, D | 1, D/2<br>2, 1-D<br>3, D/2 | 1, D<br>2, 1-D | 1, D<br>2, 1-D | 1, D<br>2, 1-D | 1, D<br>2, 1-D |
| 3 |  | 3, 1-D<br>4, D | 3, 1-D<br>4, D | 3, 1-D<br>4, D | 2, D/2<br>3, 1-D<br>4, D/2 | 2, D<br>3, 1-D | 2, D<br>3, 1-D | 2, D<br>3, 1-D |
| 4 |  | 4, 1-D<br>5, D | 4, 1-D<br>5, D | 4, 1-D<br>5, D | 4, 1-D<br>5, D | 3, D/2<br>4, 1-D<br>5, D/2 | 3, D<br>4, 1-D | 3, D<br>4, 1-D |
| 5 |  | 5, 1-D<br>6, D | 5, 1-D<br>6, D | 5, 1-D<br>6, D | 5, 1-D<br>6, D | 5, 1-D<br>6, D | 4, D/2<br>5, 1-D<br>6, D/2 | 4, D<br>5, 1-D |
| 6 |  | 6, 1 | 6, 1 | 6, 1 | 6, 1 | 6, 1 | 6, 1 | 5, D<br>6, 1-D |

**Table S2.** Drift Matrix for Model 2

| | $R_{j^*}$ | 0 | 1 | 2 | 3 | 4 | 5 | 6 |
| --- | --- | --- | --- | --- | --- | --- | --- | --- |
| $I_h$ | | | | | | | | |
| 0 |  | 0, 1-D<br>1, D | 0, 1 | 0, 1 | 0, 1 | 0, 1 | 0,1 | 0,1 |
| 1 |  | 1, 1-D<br>2, D | 0, D/2<br>1, 1-D<br>2, D/2 | 0, D<br>1, 1-D | 1,1 | 1,1 | 1,1 | 1,1 |
| 2 |  | 2,1 | 2, 1-D<br>3, D | 1, D/2<br>2, 1-D<br>3, D/2 | 1, D<br>2, 1-D | 2,1 | 2,1 | 2,1 |
| 3 |  | 3,1 | 3,1 | 3, 1-D<br>4, D | 2, D/2<br>3, 1-D<br>4, D/2 | 2, D<br>3, 1-D | 3,1 | 3,1 |
| 4 |  | 4,1 | 4,1 | 4,1 | 4, 1-D<br>5, D | 3, D/2<br>4, 1-D<br>5, D/2 | 3, D<br>4, 1-D | 4,1 |
| 5 |  | 5,1 | 5,1 | 5,1 | 5,1 | 5, 1-D<br>6, D | 4, D/2<br>5, 1-D<br>6, D/2 | 4, D<br>5, 1-D |
| 6 |  | 6, 1 | 6, 1 | 6, 1 | 6, 1 | 6, 1 | 6, 1 | 5, D<br>6, 1-D |

**Table S3.** Drift Matrix for Model 3

| | $R_{j^*}$ | 0 | 1 | 2 | 3 | 4 | 5 | 6 |
| --- | --- | --- | --- | --- | --- | --- | --- | --- |
| $I_h$ | | | | | | | | |
| 0 |  | 6, D/2<br>0, 1-D<br>1, D/2 | 6, D<br>0, 1-D | 0, 1 | 0, 1 | 0, 1 | 0,1 | 0, 1-D<br>1, D |
| 1 |  | 1, 1-D<br>2, D | 0, D/2<br>1, 1-D<br>2, D/2 | 0, D<br>1, 1-D | 1,1 | 1,1 | 1,1 | 1,1 |
| 2 |  | 2,1 | 2, 1-D<br>3, D | 1, D/2<br>2, 1-D<br>3, D/2 | 1, D<br>2, 1-D | 2,1 | 2,1 | 2,1 |
| 3 |  | 3,1 | 3,1 | 3, 1-D<br>4, D | 2, D/2<br>3, 1-D<br>4, D/2 | 2, D<br>3, 1-D | 3,1 | 3,1 |
| 4 |  | 4,1 | 4,1 | 4,1 | 4, 1-D<br>5, D | 3, D/2<br>4, 1-D<br>5, D/2 | 3, D<br>4, 1-D | 4,1 |
| 5 |  | 5,1 | 5,1 | 5,1 | 5,1 | 5, 1-D<br>6, D | 4, D/2<br>5, 1-D<br>6, D/2 | 4, D<br>5, 1-D |
| 6 |  | 5, D/2<br>6, 1-D | 6, 1 | 6, 1 | 6, 1 | 6, 1 | 6, 1-D<br>0, D/2 | 5, D/2<br>6, 1-D<br>0, D/2 |

**Table S4.**
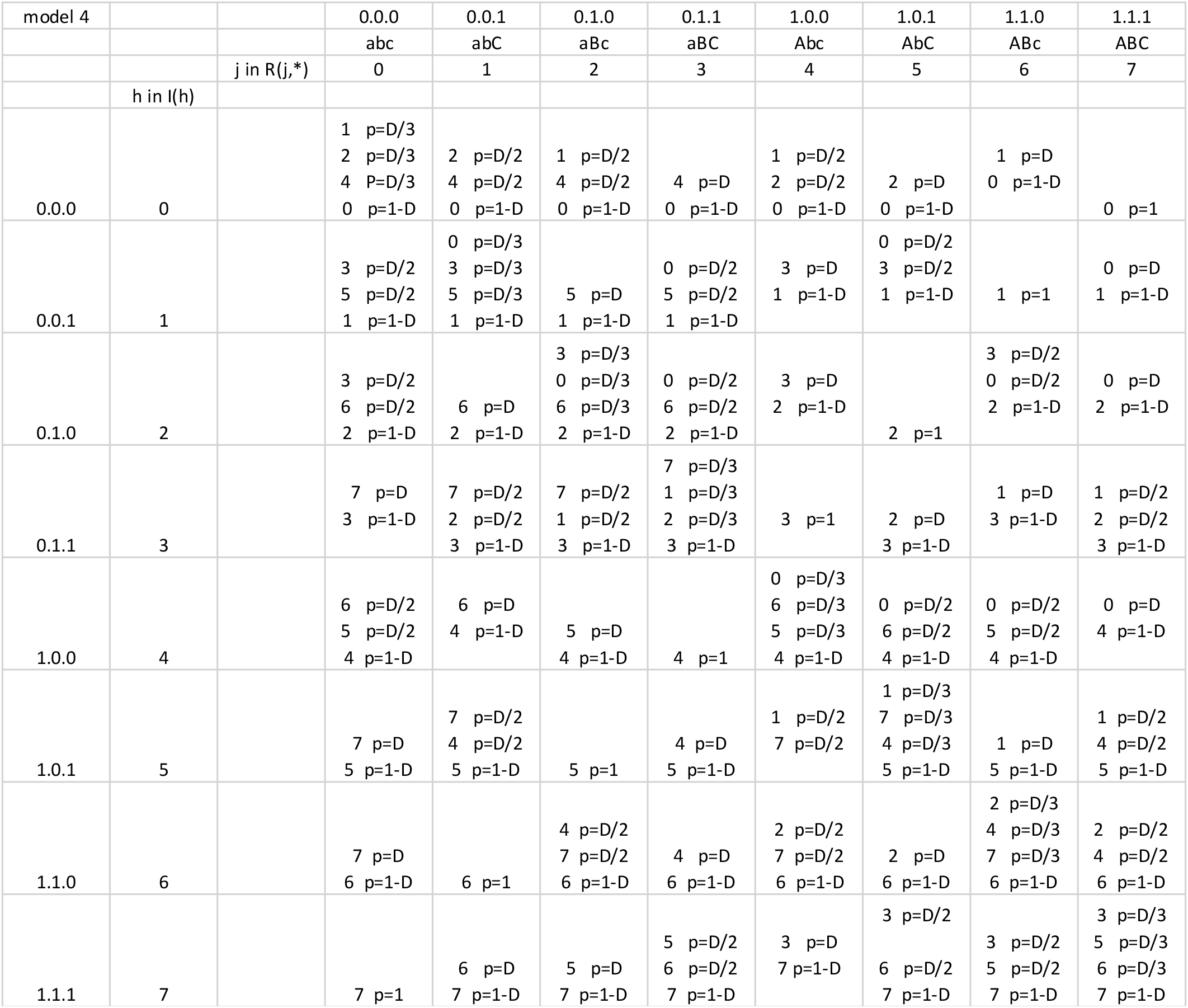
Drift Matrix for Model 4

**Figure S1.**
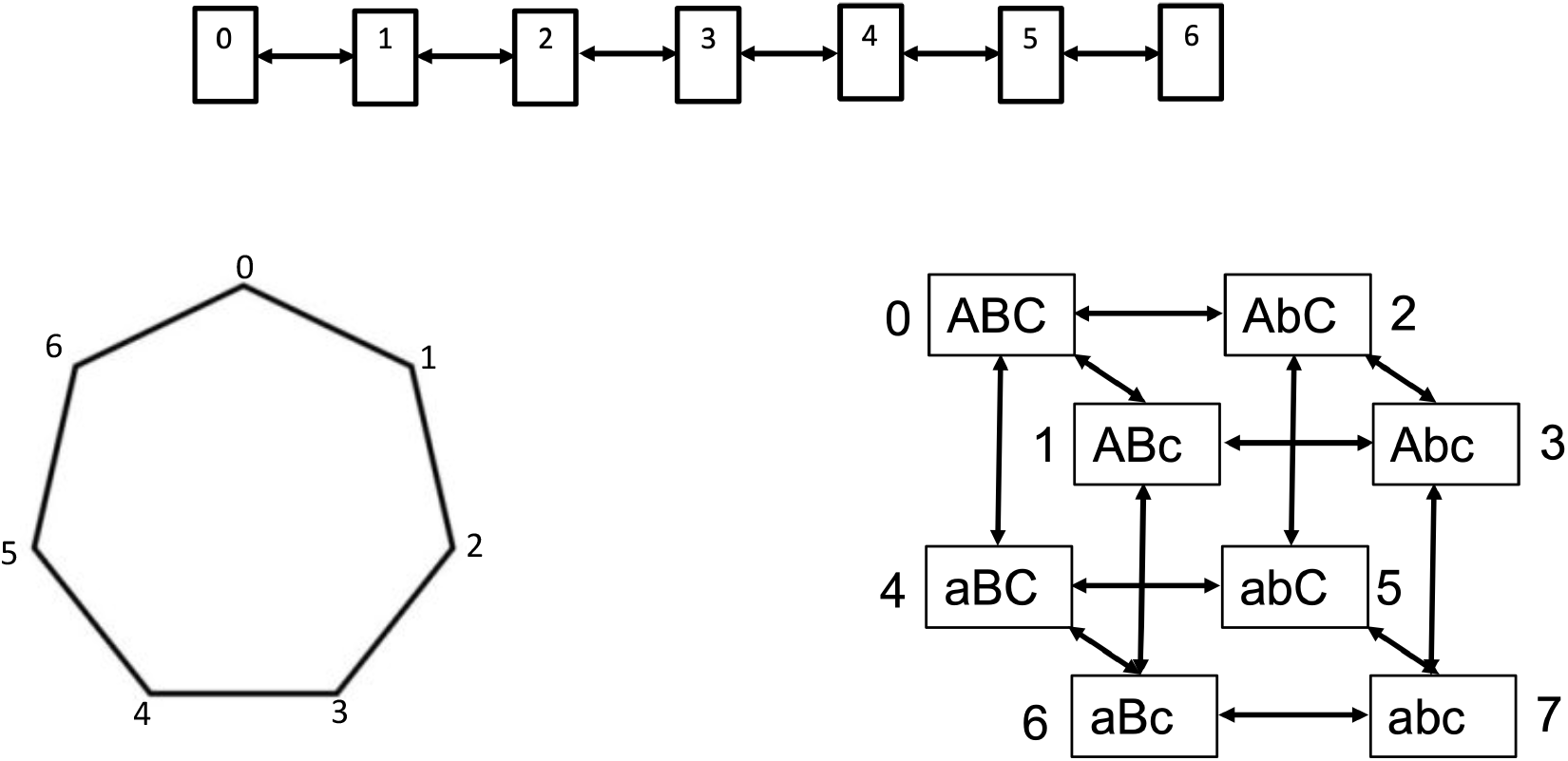
Three Model Configurations

## S2. Model Behavior Without Vaccination

### Interpreting waning rates

To provide a feel for the values of the waning parameter W, we present in Row 2 of the Table S5 below, the waning rates in terms of the time it takes for half of recently uninfected individuals to lose all of their immunity in Model 1. This is the time when half the population is at R_06_, waning level 6. In Row 3, we present the waning rates in terms of the time for the sum of all susceptibility across the whole population to reach a level that has half the immunity it had right after infection.

**Table S5.** Interpretations of the waning rates in Model 1.

| Waning Rate | 0.001 | 0.01 | 0.1 |
| --- | --- | --- | --- |
| Half Max 6 | 109 years | 11 years | 57 weeks |
| Half Popn. Immunity | 69.7 years | 7 years | 36 weeks |

Behavior of the Four Models for W=0.1 and Drifting Varies

**Figure S2.**
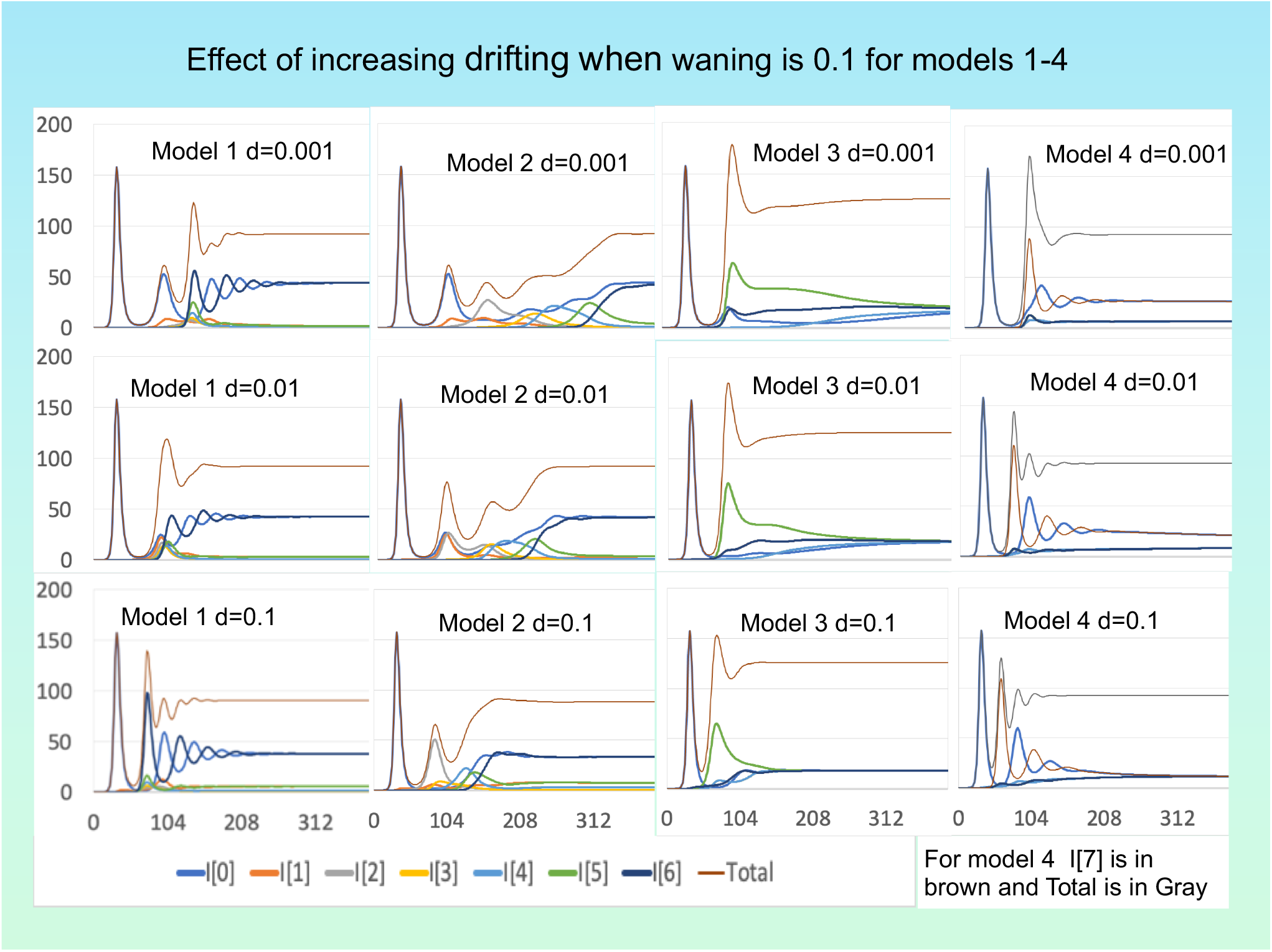
Runs of all 4 models with waning rate fixed at W=0.1 and drifting fraction varying at D=0.001, 0.01, 0.1. In each panel the top curve gives the total number of infections. Lower graphs track strains 0 through 6. (Compare to Figure 1 in the main text.)

## Model Behavior for Especially Small Values of D and W

For any W and D in or model, the first epidemic peaks around 32 weeks. When there is no waning (W=0), there are additional discrete epidemic spikes at t=4012, 6372, 8066, 9397. A similar graph holds when D=0 and W is small (0.00001), with spikes that peak a few weeks earlier, at 3990, 6336, 8020, 9342. However, with miniscule positive values of *both* W and D (W=D=0.00001), epidemic spikes (dotted curve) occur as early as t=244, 551, and 1144 weeks. It’s the interaction between waning and drifting that matters.

Figure S3 illustrates this phenomenon for Model 1. Figure S4 shows that this story persists for all four models.

**Figure S3.**
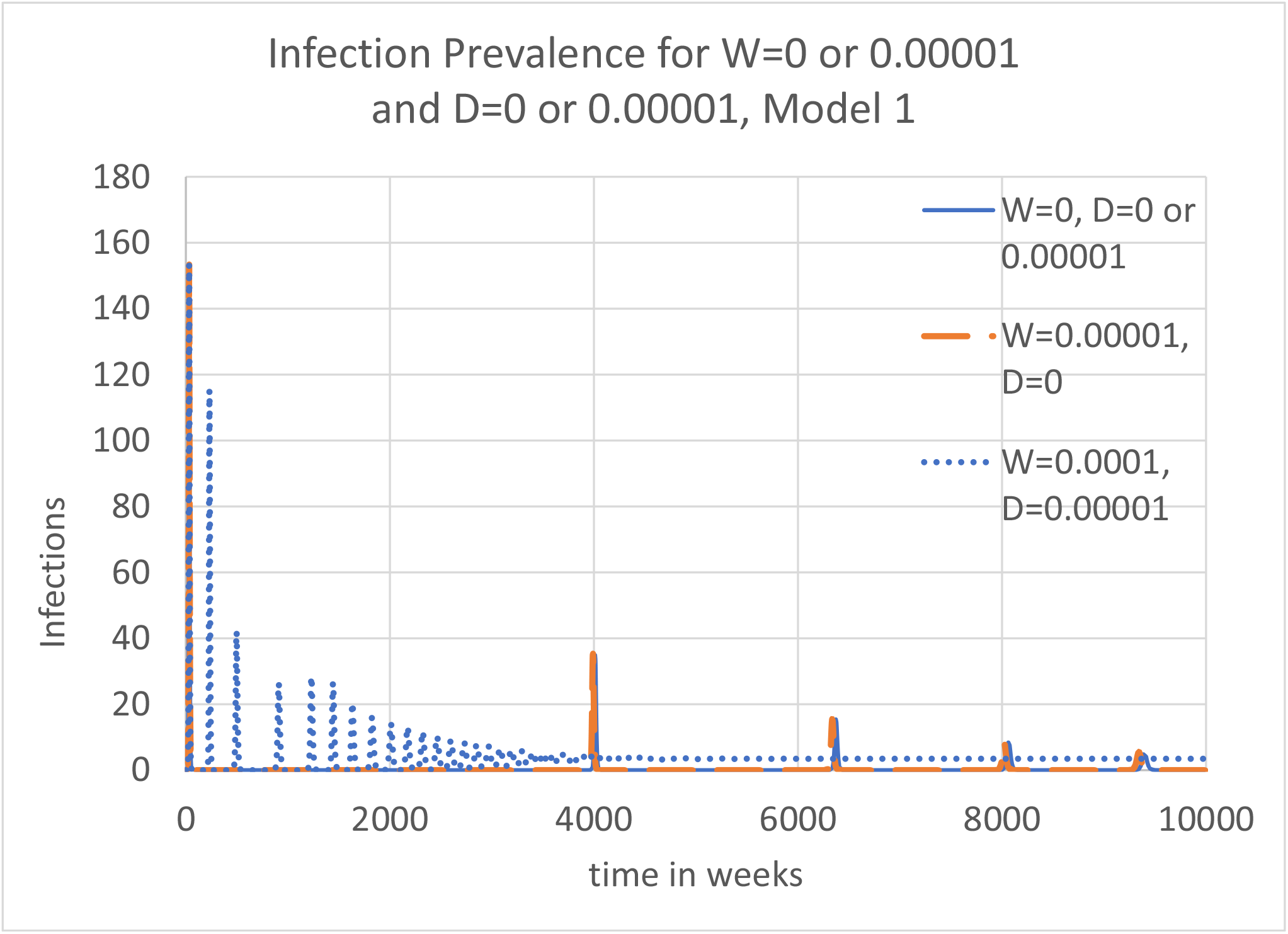
The tiniest bit of waning and drifting greatly changes infection dynamics. If W=0 and D=0.00001, the dynamics is still the same as for W=0 and D=0. If W=0.00001 and D=0, the dynamics is virtually the same. But, if W and D are <u>both</u> at the micro level 0.00001, the dynamics changes dramatically. The second epidemic now peaks 4.3 years after the first, not 75, and the next epidemic is just 6.2 years later.

**Figure S4.**
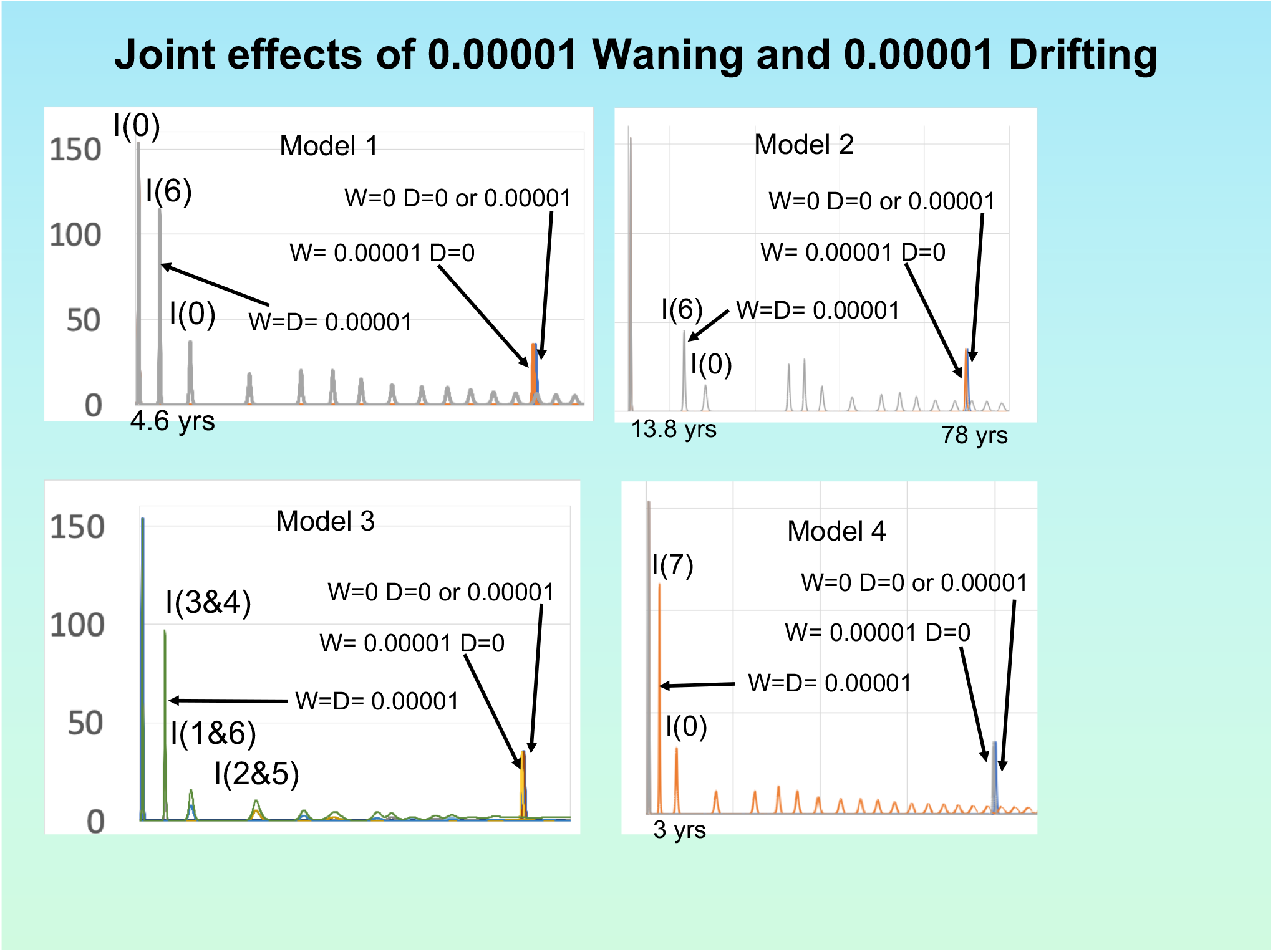
The phenomenon described in Figure S3 holds in all four models. It’s the interaction between waning and drifting that matters.

## S3. Model Behavior With Vaccination

**Figure S5.**
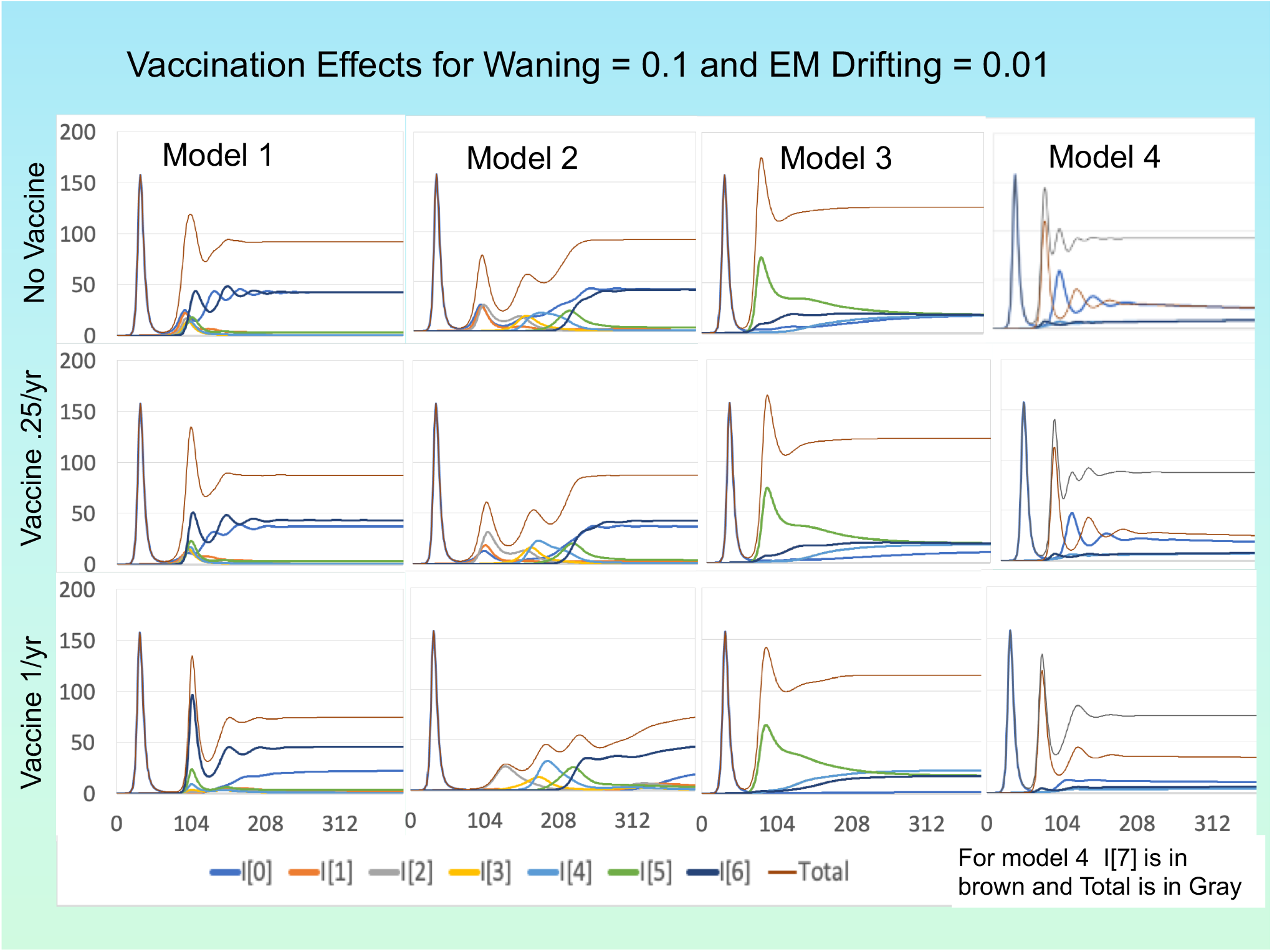
For each of the 4 models, W =0.1 and D = 0.01 are fixed. The first row presents the graphs for the case of no vaccination; the second row the case of partial vaccination (25% of the population per year); the third row full vaccination per year.

**Figure S6.**
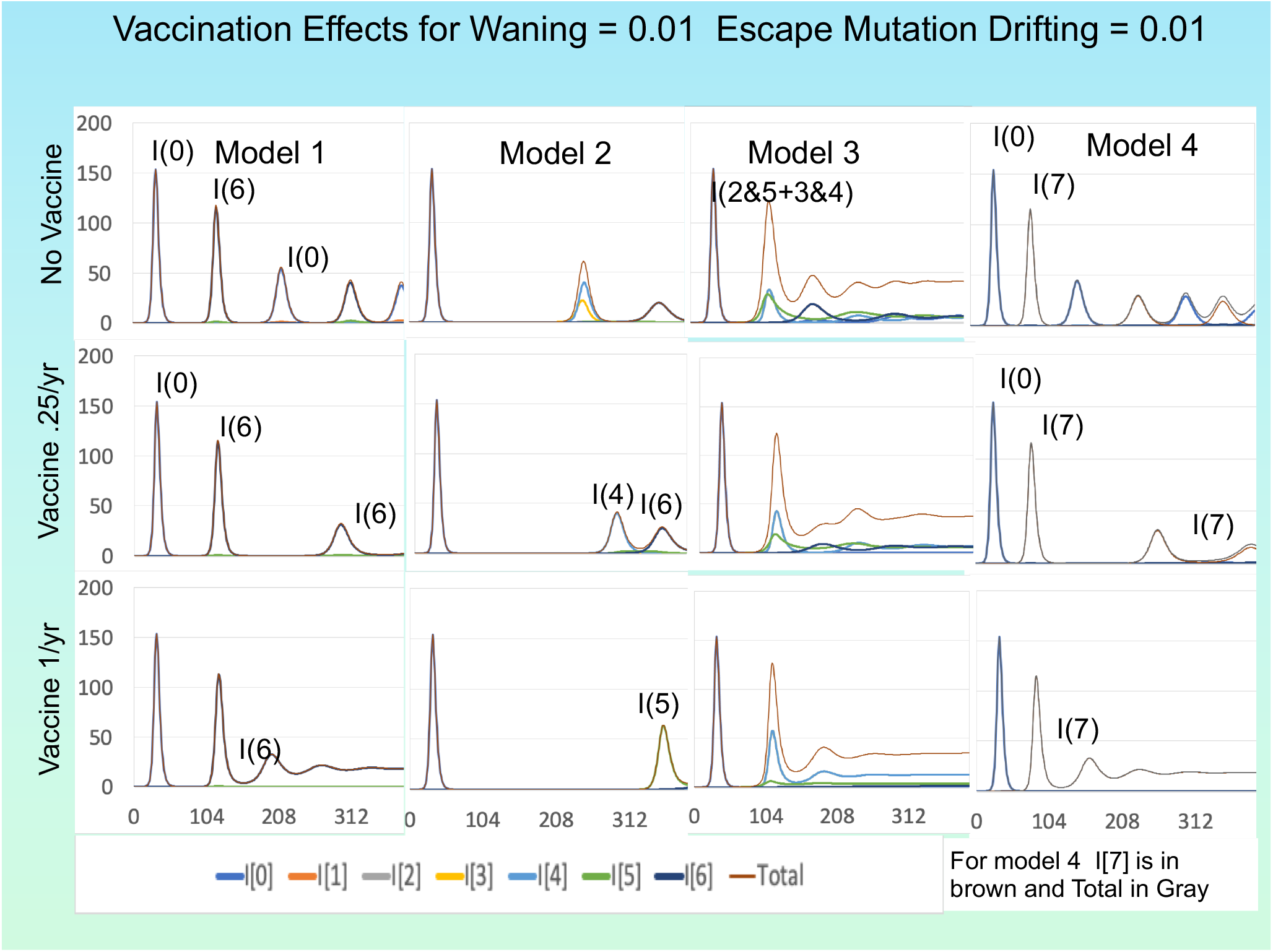
For each of the 4 models, W and D are fixed 0.01. The first row presents the graphs for the case of no vaccination; the second row the case of partial vaccination (25% of the population per year); the third row full vaccination per year.

**Figure S7.**
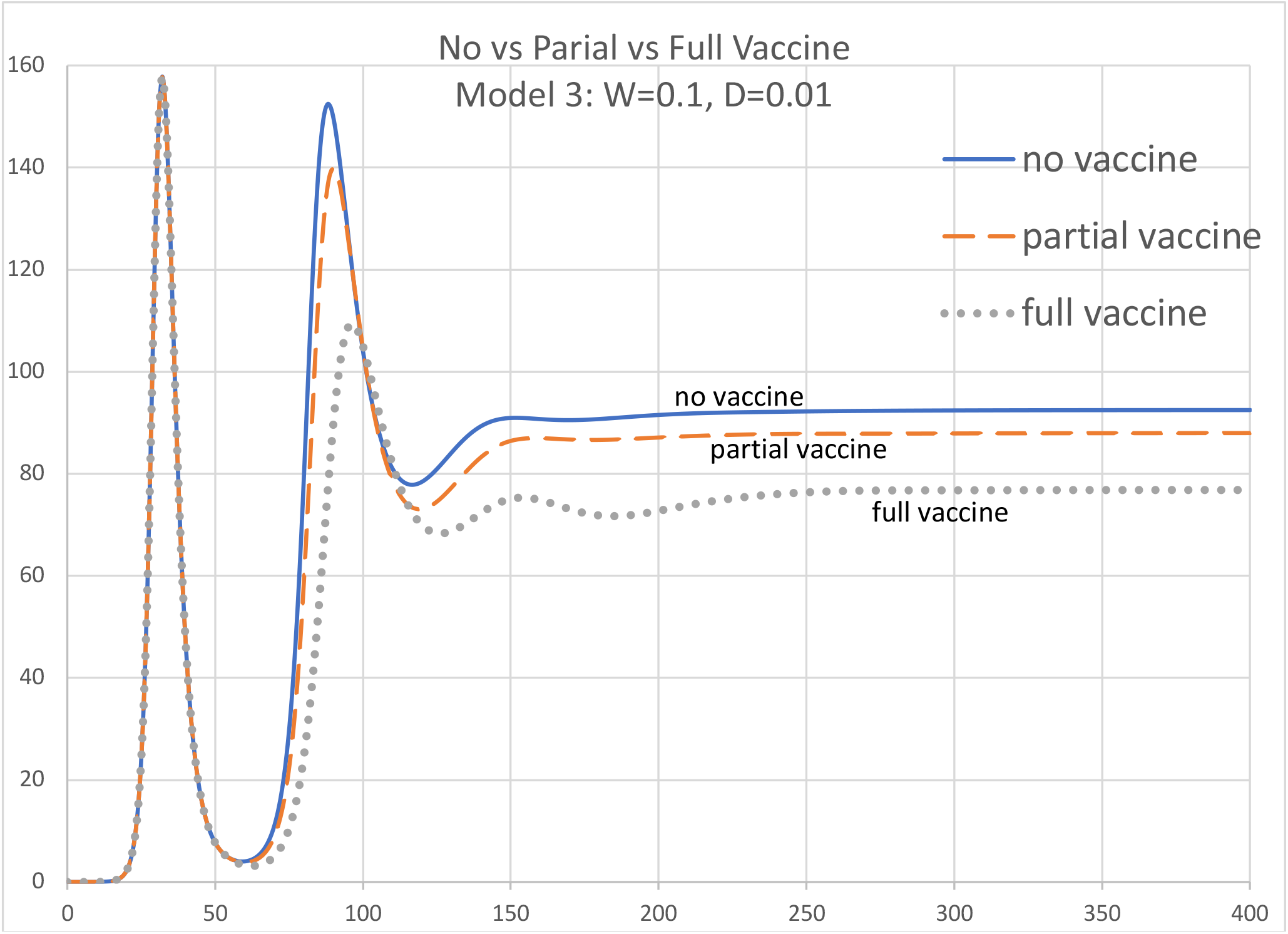
Simulation of Model 3 with W=0.1 and D=0.01 with three vaccination possibilities: none, partial (25% of the population per year, rate = 0.005/week), full (100% of the population per year, rate = 0.02/week).

**Figure S8.**
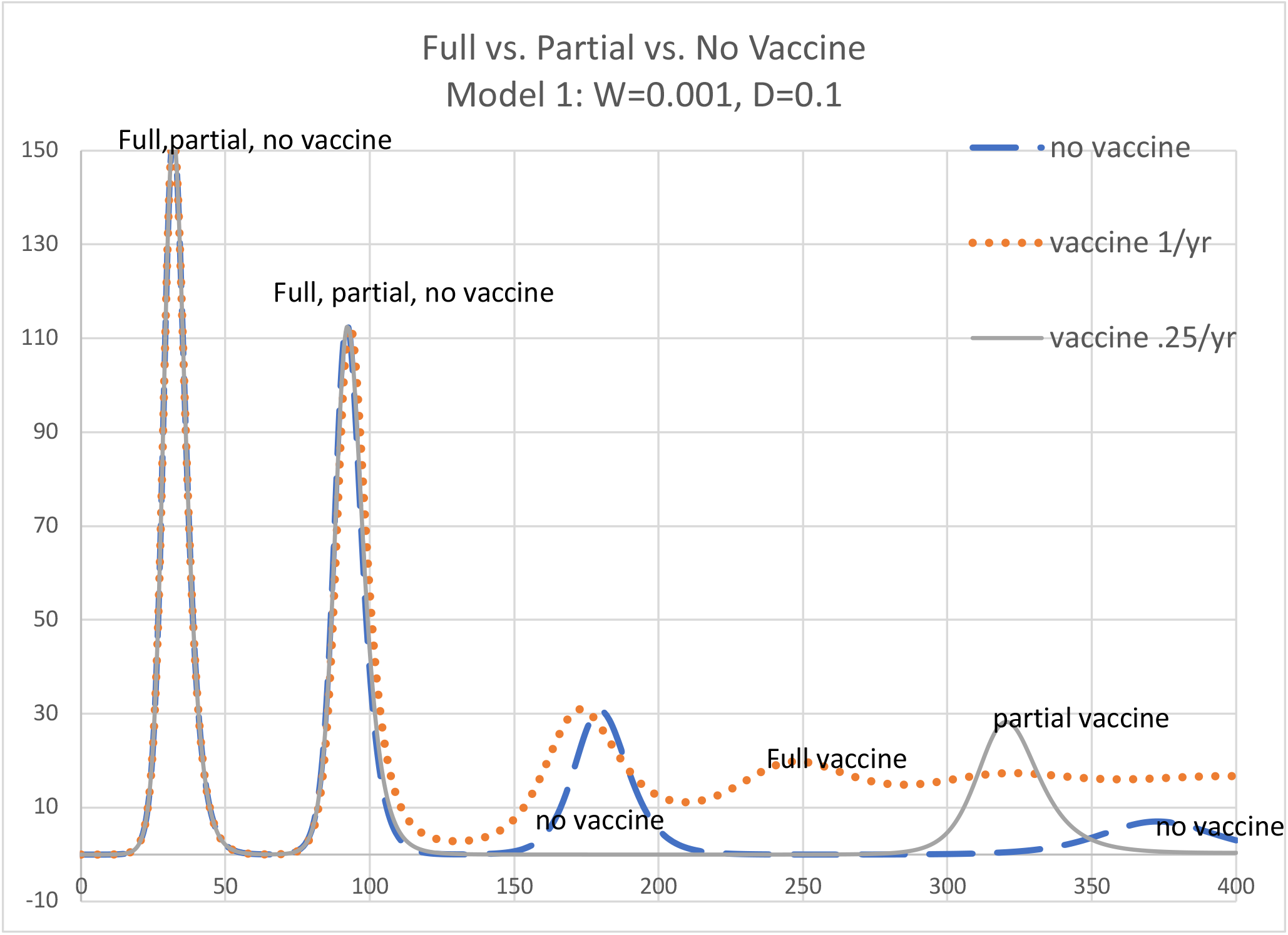
Simulation of Model 1 with W=0.001 and D=0.1 and same three vaccination possibilities as in Figure S7. Partial vaccination works well; full vaccination does not.

**Figure S9.**
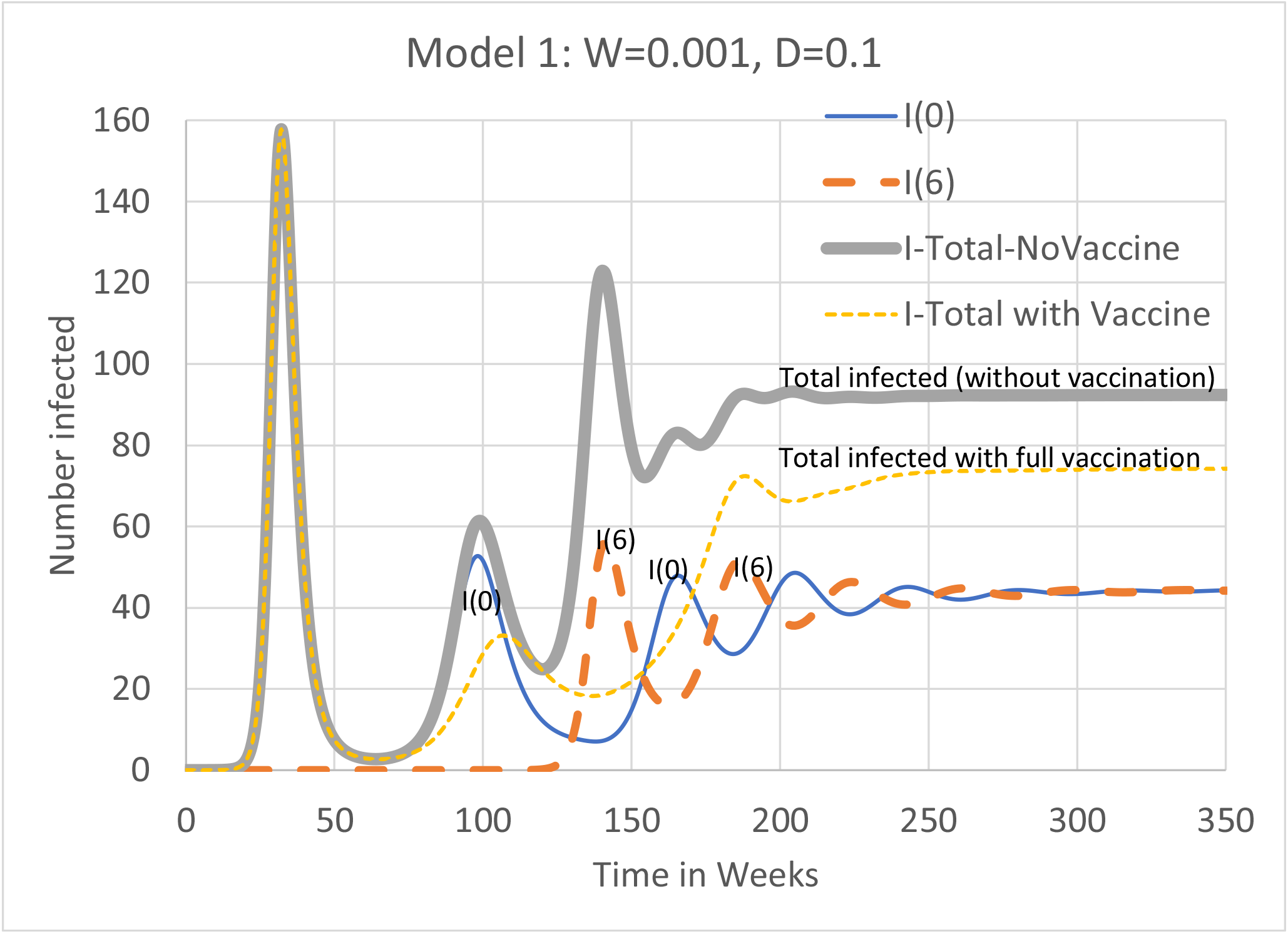
Simulation of Model 1 with W=0.001 and D=0.1, the model underlying the construction of cross-neutralization tables below. Strains 0 and 6 dominate alternate waves. Vaccination has positive effects.

## S4. Sample Computation of Model-Motivated Cross-Neutralization Tables

On this section, we continue our discussion of constructing cross-neutralization tables from our models. We work with Model 1, with waning rate W=0.001 and drifting fraction D = 0.1. This is the situation pictured in the non-vaccine graphs in Figure S9. Note the alternation of strains 0 and 6 over time, until the system reaches equilibrium.

Consider the pattern of *R*(*j,k*) individuals taken from this model output. Tables S6 and S7 presents the cross-neutralization table at week 60. The assay viruses were the original pandemic I(0) virus and the first step drift virus I(1). At time 60 almost all viruses are at I(0) so the I(1) might have been obtained elsewhere and the question is whether that virus might have drifted from the original pandemic virus. Note that the I(0) titers are all higher than the I(1) titers. This indicates that the I(1) virus has drifted. Given the high waning rate, there are highly waned immunity levels at the bottom left of the diagonal where there is not enough residual immunity to determine if there has been drifting from the I(0) virus.

**Table S6.** A cross-neutralization table generated from the *R*(*j,k*) population at week 60 given a waning rate of 0.1 per week for each of the 6 waning steps and a very small drift fraction of 0.001 per transmission.

| Drift Level 0 vs. 1; Waning rate = 0.1; drift fraction = 0.001; week = 60 |  |  |  |  |  |  |  |  |  |  |  |
| --- | --- | --- | --- | --- | --- | --- | --- | --- | --- | --- | --- |
| 10 |  |  |  |  |  |  |  |  |  | 85 |  |
| 9 |  |  |  |  |  |  |  |  | 171 |  | 0.1 |
| 8 |  |  |  |  |  |  |  | 200 |  | 0.1 |  |
| 7 |  |  |  |  |  |  |  |  | 0.1 |  |  |
| 6 |  |  |  |  |  | 166 |  | 0 | 0 |  |  |
| 5 |  |  |  |  | 107 | 0 | 0.03 | 0 |  |  |  |
| 4 |  |  |  |  | 0 | 0.01 |  |  |  |  |  |
| 3 |  |  |  | 57 | 0 |  |  |  |  |  |  |
| 2 |  |  | 41 | 0 |  |  |  |  |  |  |  |
| 1 |  |  |  |  |  |  |  |  |  |  |  |
| DL0 |  |  |  |  |  |  |  |  |  |  |  |
| virus | DL1 | 1 | 2 | 3 | 4 | 5 | 6 | 7 | 8 | 9 | 10 |

**Table S7.**
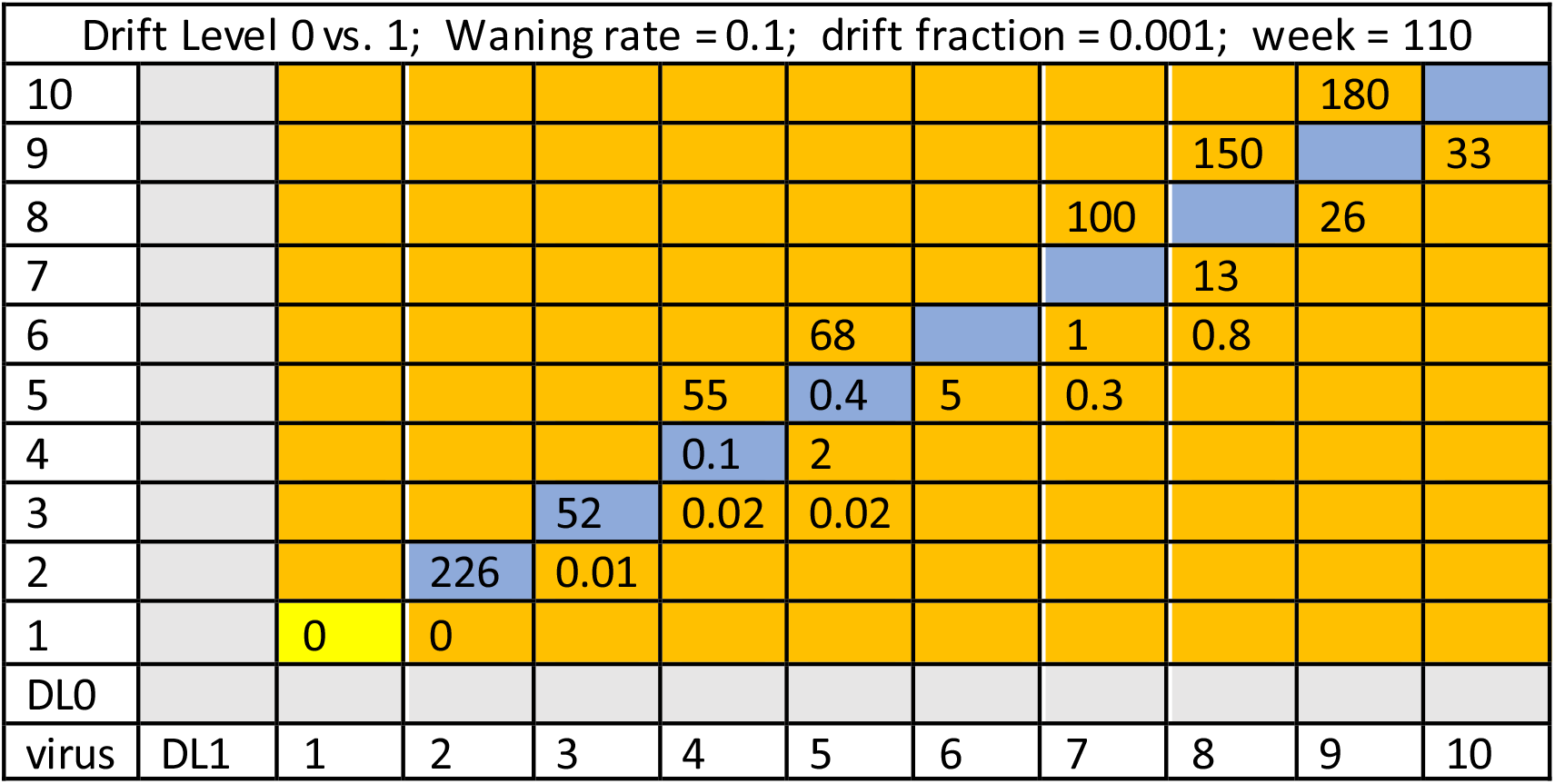
The cross-neutralization table generated from the *R*(*j,k*) population at week 110.

| Drift Level 0 vs. 1; Waning rate = 0.1; drift fraction = 0.001; week = 110 |  |  |  |  |  |  |  |  |  |  |  |
| --- | --- | --- | --- | --- | --- | --- | --- | --- | --- | --- | --- |
| 10 |  |  |  |  |  |  |  |  |  | 180 |  |
| 9 |  |  |  |  |  |  |  |  | 150 |  | 33 |
| 8 |  |  |  |  |  |  |  | 100 |  | 26 |  |
| 7 |  |  |  |  |  |  |  |  | 13 |  |  |
| 6 |  |  |  |  |  | 68 |  | 1 | 0.8 |  |  |
| 5 |  |  |  |  | 55 | 0.4 | 5 | 0.3 |  |  |  |
| 4 |  |  |  |  | 0.1 | 2 |  |  |  |  |  |
| 3 |  |  |  | 52 | 0.02 | 0.02 |  |  |  |  |  |
| 2 |  |  | 226 | 0.01 |  |  |  |  |  |  |  |
| 1 |  | 0 | 0 |  |  |  |  |  |  |  |  |
| DL0 |  |  |  |  |  |  |  |  |  |  |  |
| virus | DL1 | 1 | 2 | 3 | 4 | 5 | 6 | 7 | 8 | 9 | 10 |

Next look at the cross-neutralization pattern at 110 weeks in Table S7. At that time there are clearly enough individuals whose I(1) neutralization titer is higher than their I(0) titer to conclude that drifted viruses have circulated in this population. As seen in Figure S9, 110 weeks is well into the second wave of the epidemic but before the I(6) viruses surge to co-dominate the epidemic along with the I(0) strain. At 110 weeks, however, individuals whose major immunity is to an I(0) virus are more than 13 times as numerous as individuals whose major immunity is to a drifted virus beyond I(0). Thus this population is set up for rapid drifting since all viruses that have a drifting level to the right of I(0) have escape mutations to I(0). Consequently these virus levels will have a higher rate of infection than I(0) viruses and will experience a strong selective force to drive further drifting. The realization of that drifting pressure is evident in Figure 4. That high degree of drifting occurred even though the drifting fraction parameter is very low.

When vaccination at a rate of 0.02 per week (more than once a year) begins on week 52, we see in Figure S9 that for these parameter values there is no noticeable infection or virus drifting until about week 120. At t=120, infections with drifted viruses rise to a prevalence of 2/1000. Thereafter the epidemic is almost as explosive as the first epidemic. That is because 99.9% of the population has 6/7 of the susceptibility to the rising I(6) virus as the original population had to the I(0) pandemic strain.

If the world experiences escape mutations as intensely as our model, there won’t be time to develop vaccines that address these escape mutations. But we suspect that when the simplifying assumptions we listed earlier are addressed, the intensity of escape mutations will diminish.

## S5. Changing other parameters

We changed various parameters in our basic model, both to understand their roles and to ascertain our model’s robustness. In particular, we varied 1) the strain on which the vaccine focused, 2) the overall contagiousness of the infection and consequently the underlying R_0_, 3) the relative effect of drifting on contagiousness in expression (1.1), and 4) the relative effect of waning on contagiousness in (1.1).

**Figure S10.**
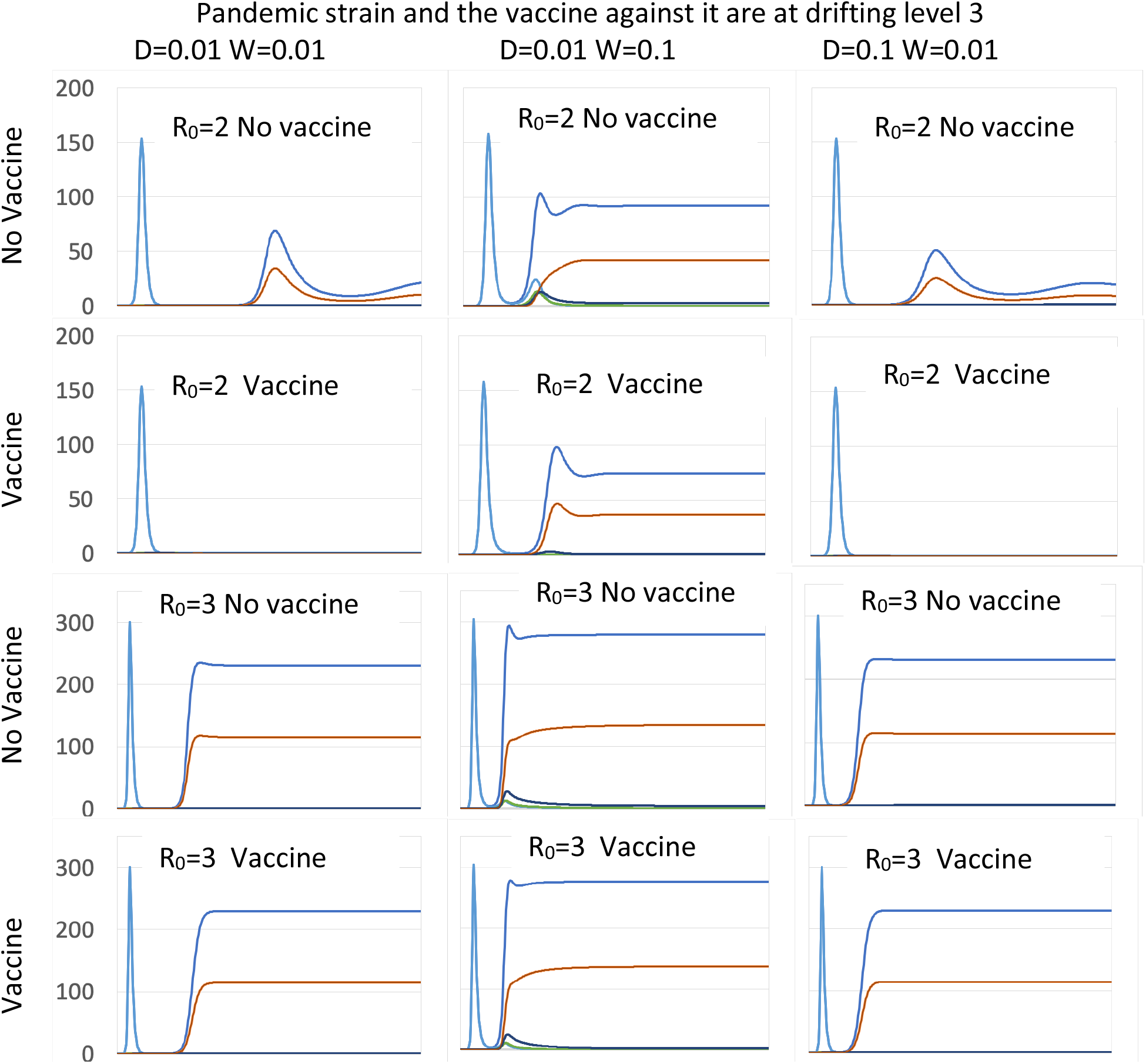
Simulation of Model 1 where Strain 3 is the original strain and vaccine focus. In the top two rows, R_0_=2 (as in most of this paper). The vaccine works well when W=0.001, but not so well when W=0.1. R_0_ is increased to 3 in the bottom two rows; vaccine is ineffective at all values of W.

When the initial strain is I_0_ and the vaccine focuses on that strain, for low values of W the vaccine can do more harm than good. But what if the initial/focal strain is not *I*_*0*_? By the symmetry of our model, similar results would hold if we replaced *I*_*0*_ by *I*_*6*_. A more interesting case is allowing the initial level to be the internal level *I*_*3*_ with a vaccine moving susceptibles to *R*_*30*_. The first two rows in Figure S10 illustrate the positive effects of the vaccine, depending on the waning rate W. If W is as high as 0.1, then the vaccine reduces the prevalence of the infection after the first wave. This is shown in the second column of Figure S10 for D=0.01, but holds more generally. If W is at a lower level of 0.01, then the vaccine can postpone rebound epidemics for long times after the first infection (columns 1 and 3, rows 1 and 2 in Figure S10).

However, all the simulations up to this point were run under the assumption that the basic reproduction number R_0_ is 2. If instead R_0_ = 3, then as the last two rows of Figure S10 illustrate, the vaccine has virtually no effect on infection prevalence.

Finally, we examine the robustness of our conclusion to changes in the parameters in the expression (1.1) for probability of transmission. Our simulations used Q=P+1 and L=M in (1.1). If we increase Q, we are reducing the relative effect that drifting has on transmission; if we increase L, we are reducing the relative impact of waning. Figure 7 illustrates the effect of such changes on vaccine success. In the first column, we have reduced the transmission potential of drifting (TPD) by 80% by changing Q from (P+1) to 5(P+1) in (1.1). In the second column, we have reduced the transmission potential of waning (TPW) by 80% by changing L from M to 5M in (1.1). We remain in the situation of the lower half of Figure S10 where *R*_*0*_ = 3 and the initial drift level and vaccine target is level 3, the situation in which the vaccine had little effect. Figure S11 shows in this case that there is still very little effect when the TPW is dramatically lower. However, given an 80% reduction in drifting effect on increasing susceptibility to reinfection, the vaccine can have a noticeable effect on the epidemic after the first wave.

**Figure S11.**
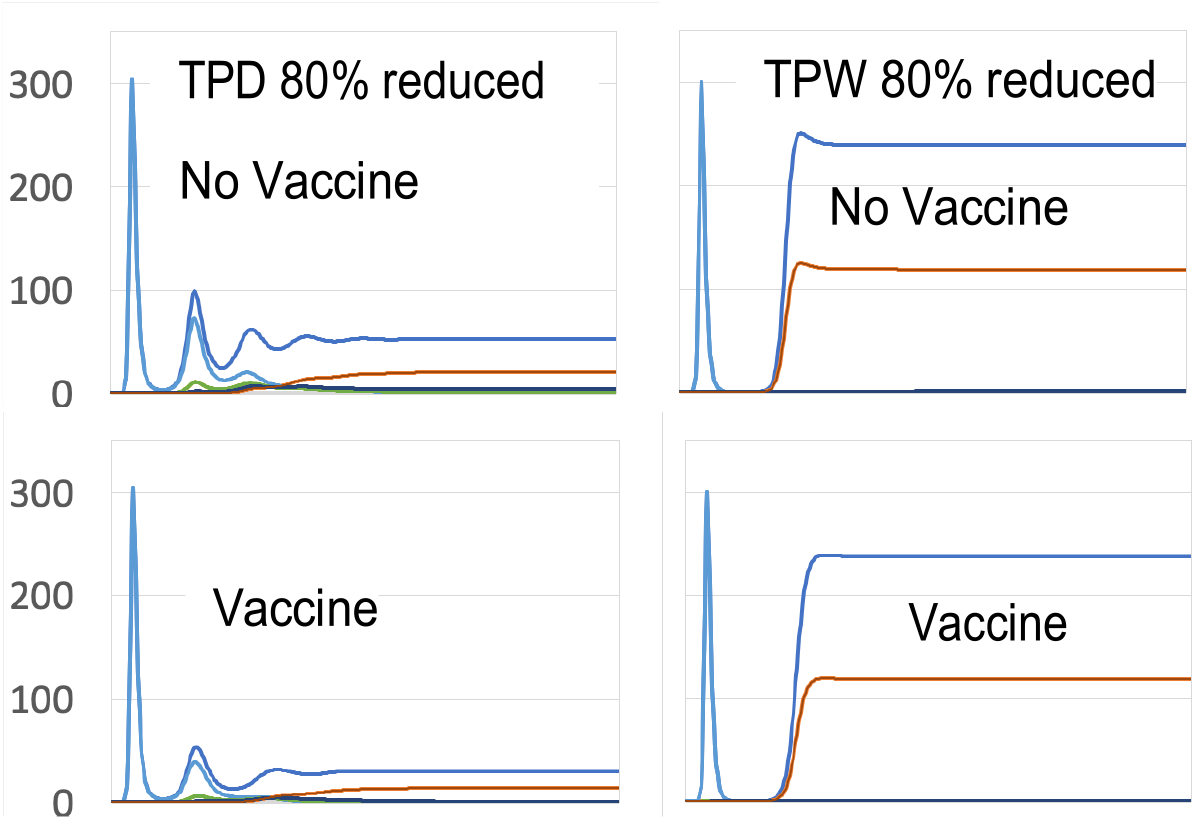
Simulation of Model 1 with D=0.01, W=0.1, and R_0_=3, as in the last two rows of the middle column in Figure S10. In the first column, the effect of *drifting* on transmission function Z is reduced 80%: as a result, prevalence is reduced, especially after vaccination. In the second column, the effect of *waning* on transmission Z is reduced 80%: no effect before or after vaccination.

In an epidemic that includes waning immunity and antigenic drifting to escape such immunity, we need a systems approach to understand the dynamics of the infection in order to gauge the possible effects of a vaccine.

## S6. Paths to epitope data informed vaccine decisions using DRIA

A wide variety of scientists are exploring epitope contributions to immunity. The determination of epitope location is informed by

- model based and artificial intelligence enhanced protein folding analyses,
- linear epitope mapping using CRISPR enhanced construction of epitopes and epitope variants,
- serological assays using epitope specific antigens,
- in silico analyses of proteins to identify sites where B and T cell antigens might be processed by different host genetic states,
- molecular epidemiology based serological analyses of immune responses,
- evolutionary biology models and analyses, and epidemiological model fitting.

The scientists using these approaches have a wide variety of philosophies of science guiding their search for new knowledge. Yet their work must be integrated if epitope specified models of host immunity and viral evolution are to be successful. This will not be an easy task.

Scientists in these specialties naturally want to pour the knowledge and methods available in their specialties into uncovering new knowledge about SARS-CoV-2. Epidemiologists who want to guide public health decisions need a context for integrating all the vast new knowledge that is being generated. A model fitted to data provides a particular context. (Davies, Barnard et al. 2021) have fitted a model to SARS-CoV-2 data that is strain specific and could inform Public Health decisions regarding the new B.1.1.7 variant. They have added much realistic complexity to their model that is relevant to public health decisions. (Volz, Mishra et al. 2020) have fit epidemiological models to the new B.1.1.7 using an approach that uses more extensive location-specific model fits to make inferences about virus variants on transmission dynamics. That approach seems to use the data more fully. The estimates of increased transmissibility of B.1.1.7 from the two groups are about the same.

How these analyses can inform decisions about changes in vaccine composition to counteract escape mutations is not clear. We believe that a DRIA approach can enable models to use epitope specific immunological data like that generated by (Greaney, Starr et al. 2020, Greaney, Loes et al. 2021) and (Shrock, Fujimura et al. 2020). Cross-neutralization assays could play a key role.

One way to start is to fit cross-neutralization assays to the extensive populations that are under study in vaccine trials. In addition, extensive populations in serological surveys could provide key data. The unit of analysis in such studies is the individual. At this level the joint effects of different antibodies to different epitopes could be described. Such models could then be incorporated directly into the type of models we have presented here. There then could be a second stage of fitting performed in the DRIA context to inform decisions about epitope composition of vaccines.

## REFERENCES

Andersen, K. G., A. Rambaut, W. I. Lipkin, E. C. Holmes and R. F. Garry (2020). “The proximal origin of SARS-CoV-2.” Nat Med 26(4): 450–452.

Andreasen, V., J. Lin and S. A. Levin (1997). “The dynamics of cocirculating influenza strains conferring partial cross-immunity.” J Math Biol 35(7): 825–842.

Babiker, A., C. Marvil, J. J. Waggoner, M. Collins and A. Piantadosi (2020). “The Importance and Challenges of Identifying SARS-CoV-2 Reinfections.” J Clin Microbiol.

Davies, N. G., R. C. Barnard, C. I. Jarvis, A. J. Kucharski, J. Munday, C. A. B. Pearson, T. W. Russell, D. C. Tully, S. Abbott, A. Gimma, W. Waites, K. L. M. Wkong, K. van Zandvoort, … and W. J. Edmonds (2021). “Estimated transmissibility and severity of novel SARS-CoV-2 Variant of Concern 202012/01 in England.” medRxiv.

Eguia, R., K. H. D. Crawford, L. Kelnhofer-Milevolte, A. L. Greninger, J. A. Englund, M. J. Boeckh and J. D. BLoom (2020). “A human coronavirus evolves antigenically to escape antibody immunity.” bioRxiv.

Funk, S. and A. A. King (2019). “Choices and trade-offs in inference with infectious disease models.” Epidemics 30: 100383.

Greaney, A. J., A. N. Loes, K. H. D. Crawford, T. N. Starr, K. D. Malone, H. Y. Chu and J. D. Bloom (2021). “Comprehensive mapping of mutations to the SARS-CoV-2 receptor-binding domain that affect recognition by polyclonal human serum antibodies.” bioRxiv.

Greaney, A. J., T. N. Starr, P. Gilchuck, S. J. Zost, E. Binshtein, S. K. Hilton, J. Huddleston, R. Eguia, K. H. D. Crawford, K. D. Malone, …, J. E. Crowe Jr., and J. D. Bloom (2020). “Complete Mapping of Mutations to the SARS-CoV-2 Spike Receptor-Binding Domain that Escape Antibody Recognition.”

Harrington, D., B. Kele, S. Pereira, X. Couto-Parada, A. Riddell, S. Forbes, H. Dobbie and T. Cutino-Moguel (2021). “Confirmed Reinfection with SARS-CoV-2 Variant VOC-202012/01.” Clin Infect Dis.

Hou, Y. J., S. Chiba, P. Halfmann, C. Ehre, M. Kuroda, K. H. Dinnon, 3rd, S. R. Leist, A. Schafer, N. Nakajima, K. Takahashi, R. E. Lee, T. M. Mascenik, R. Graham, C. E. Edwards, L. V. Tse, K. Okuda, A. J. Markmann, L. Bartelt, A. de Silva, D. M. Margolis, R. C. Boucher, S. H. Randell, T. Suzuki, L. E. Gralinski, Y. Kawaoka and R. S. Baric (2020). “SARS-CoV-2 D614G variant exhibits efficient replication ex vivo and transmission in vivo.” Science 370(6523): 1464–1468.

Kistler, K. E. and T. Bedford (2020). “Evidence for adaptive evolution in the receptor-binding domain of seasonal coronaviruses.” bioRxiv.

Lau, S. K., P. Lee, A. K. Tsang, C. C. Yip, H. Tse, R. A. Lee, L. Y. So, Y. L. Lau, K. H. Chan, P. C. Woo and K. Y. Yuen (2011). “Molecular epidemiology of human coronavirus OC43 reveals evolution of different genotypes over time and recent emergence of a novel genotype due to natural recombination.” J Virol 85(21): 11325–11337.

Madonna, B. (2021).from https://berkeley-madonna.myshopify.com/.

Monto, A. S., P. M. DeJonge, A. P. Callear, L. A. Bazzi, S. B. Capriola, R. E. Malosh, E. T. Martin and J. G. Petrie (2020). “Coronavirus Occurrence and Transmission Over 8 Years in the HIVE Cohort of Households in Michigan.” J Infect Dis 222(1): 9–16.

Nickbakhsh, S., A. Ho, D. F. P. Marques, J. McMenamin, R. N. Gunson and P. R. Murcia (2020). “Epidemiology of Seasonal Coronaviruses: Establishing the Context for the Emergence of Coronavirus Disease 2019.” J Infect Dis 222(1): 17–25.

Rambaut, A., N. J. Loman, O. G. Pybus, W. Barclay, j. Barrett, A. Carabelli, T. R. Connor, T. Peacock, D. L. Robertson and E. Volz. (2020). “Preliminary genomic characterisation of an emergent SARS-CoV-2 lineage in the UK defined by a novel set of spike mutations.” COVID-19 Genomics Consortium UK from https://virological.org/t/preliminary-genomic-characterisation-of-an-emergent-sars-cov-2-lineage-in-the-uk-defined-by-a-novel-set-of-spike-mutations/563.

Saad-Roy, C. M., C. E. Wagner, R. E. Baker, S. E. Morris, J. Farrar, A. L. Graham, S. A. Levin, M. J. Mina, C. J. E. Metcalf and B. T. Grenfell (2020). “Immune life history, vaccination, and the dynamics of SARS-CoV-2 over the next 5 years.” Science 370(6518): 811–818.

Shrock, E., E. Fujimura, T. Kula, R. T. Timms, I. H. Lee, Y. Leng, M. L. Robinson, B. M. Sie, M. Z. Li, Y. Chen, J. Logue, A. Zuiani, D. McCulloch, F. J.N. Lelis, S. Henson, D. R. Monaco, M. Travers, S. Habibi, W. A. Clarke, P. Caturegli, O. Laeyendecker, A. Piechocka-Trocha, J. Z. Li, A. Khatri, H. Y. Chu, M. C.-. Collection, T. Processing, A. C. Villani, K. Kays, M. B. Goldberg, N. Hacohen, M. R. Filbin, X. G. Yu, B. D. Walker, D. R. Wesemann, H. B. Larman, J. A. Lederer and S. J. Elledge (2020). “Viral epitope profiling of COVID-19 patients reveals cross-reactivity and correlates of severity.” Science 370(6520).

Tegally, H., E. Wilkinson, M. Giovanetti, A. Iranzadeh, V. Fonesca and T. Oliviera (2021). “Emergence and rapid spread of a new severe acute respiratory syndrome-related coronavirus 2 (SARS-CoV-2) lineage with multiple spike mutations in South Africa.” MedRxiv.

Volz, E., V. Hill, J. T. McCrone, A. Price, D. Jorgensen, A. O’Toole, J. Southgate, R. Johnson, B. Jackson, F. F. Nascimento, S. M. Rey, S. M. Nicholls, R. M. Colquhoun, A. da Silva Filipe, J. Shepherd, D. J. Pascall, R. Shah, N. Jesudason, K. Li, R. Jarrett, N. Pacchiarini, M. Bull, L. Geidelberg, I. Siveroni, C.-U. Consortium, I. Goodfellow, N. J. Loman, O. G. Pybus, D. L. Robertson, E. C. Thomson, A. Rambaut and T. R. Connor (2021). “Evaluating the Effects of SARS-CoV-2 Spike Mutation D614G on Transmissibility and Pathogenicity.” Cell 184(1): 64–75 e11.

Volz, E., S. Mishra, M. Chand, B. J.C.R. Johnson, L. Geidelberg, W. R. Hinsley, D. J. Laydon, D.G.A. O’Toole, A.R.M. Ragonnet-Cronin, I. Harrison, B. Jackson, C. V. Arianni, O. Boyd, N. J. Loman, J. T. McCrone, S. Goncalves, D. Jorgensen, R. Myers, V. Hill, D. K. Jackson, K. Gaythorpe, N. Groves, J. Silltoe, D. P. Kwiatkowski, S. Flaxman, O. Ratmann, S. Bhatt, S. Hopkins, A. Gamdy, A. Rambaut and N. Ferguson (2020). “Transmission of SARS-CoV-2 Lineage B.1.1.7 in England: Insights from linking epidemiological and genetic data.” bioRxiv.

Weisblum, Y., F. Schmidt, F. Zhang, J. DaSilva, D. Poston, J. C. C. Lorenzi, F. Muecksch, M. Rutkowska, H. H. Hoffmann, E. Michailidis, C. Gaebler, M. Agudelo, A. Cho, Z. Wang, A. Gazumyan, M. Cipolla, L. Luchsinger, C. D. Hillyer, M. Caskey, D. F. Robbiani, C. M. Rice, M. C. Nussenzweig, T. Hatziioannou and P. D. Bieniasz (2020). “Escape from neutralizing antibodies by SARS-CoV-2 spike protein variants.” bioRxiv.

Weissman, D., M. G. Alameh, T. de Silva, P. Collini, H. Hornsby, R. Brown, C. C. LaBranche, R. J. Edwards, L. Sutherland, S. Santra, K. Mansouri, S. Gobeil, C. McDanal, N. Pardi, N. Hengartner, P. J. C. Lin, Y. Tam, P. A. Shaw, M. G. Lewis, C. Boesler, U. Sahin, P. Acharya, B. F. Haynes, B. Korber and D. C. Montefiori (2020). “D614G Spike Mutation Increases SARS CoV-2 Susceptibility to Neutralization.” Cell Host Microbe.

Zhang, Y., J. Li, Y. Xiao, J. Zhang, Y. Wang, L. Chen, G. Paranhos-Baccala, L. Ren and J. Wang (2015). “Genotype shift in human coronavirus OC43 and emergence of a novel genotype by natural recombination.” J Infect 70(6): 641–650.

Zhu, Y., C. Li, L. Chen, B. Xu, Y. Zhou, L. Cao, Y. Shang, Z. Fu, A. Chen, L. Deng, Y. Bao, Y. Sun, L. Ning, C. Liu, J. Yin, Z. Xie and K. Shen (2018). “A novel human coronavirus OC43 genotype detected in mainland China.” Emerg Microbes Infect 7(1): 173.

## References

Shrock, E., E. Fujimura, T. Kula, R. T. Timms, I. H. Lee, Y. Leng, M. L. Robinson, B. M. Sie, M. Z. Li, Y. Chen, J. Logue, A. Zuiani, D. McCulloch, F. J. N. Lelis, S. Henson, D. R. Monaco, M. Travers, S. Habibi, W. A. Clarke, P. Caturegli, O. Laeyendecker, A. Piechocka-Trocha, J. Z. Li, A. Khatri, H. Y. Chu, M. C.-. Collection, T. Processing, A. C. Villani, K. Kays, M. B. Goldberg, N. Hacohen, M. R. Filbin, X. G. Yu, B. D. Walker, D. R. Wesemann, H. B. Larman, J. A. Lederer and S. J. Elledge (2020). “Viral epitope profiling of COVID-19 patients reveals cross-reactivity and correlates of severity.” Science 370(6520).

Volz, E., S. Mishra, M. Chand, B. J.C. R. Johnson, L. Geidelberg, W. R. Hinsley, D. J. Laydon, D. G.A. O’Toole, A. R., M. Ragonnet-Cronin, I. Harrison, B. Jackson, C. V. Arianni, O. Boyd, N. J. Loman, J. T. McCrone, S. Goncalves, D. Jorgensen, R. Myers, V. Hill, D. K. Jackson, K. Gaythorpe, N. Groves, J. Silltoe, D. P. Kwiatkowski, S. Flaxman, O. Ratmann, S. Bhatt, S. Hopkins, A. Gamdy, A. Rambaut and N. Ferguson (2020). “Transmission of SARS-CoV-2 Lineage B.1.1.7 in England: Insights from linking epidemiological and genetic data.” bioRxiv.

